# Institutionalizing LLM-assisted decision support for malaria risk-focused ITN reprioritization in Nigeria: Digital competency, workplace resource profiles, experiences, and pathways to routine integration

**DOI:** 10.64898/2026.08.19.26360823

**Authors:** Laurette Mhlanga, Bernard O. Boateng, Eniola A. Bamgboye, Hephzibah A.T. Adenji, Grace Legris, Yusuf Jamiu, Gift Enang, Maikore K. Ibrahim, Chukwu Okoronkwo, Ifeoma D. Ozodiegwu

## Abstract

**Background:** In Nigeria, the country with the greatest global malaria burden, funding constraints increasingly require insecticide-treated net (ITN) reprioritization to target those at highest risk. Large language models (LLM) assisted decision-support tools may facilitate risk-informed ITN planning by supporting malaria programme officers in navigating analyses, interpreting outputs, and translating evidence into operational decisions. We developed ChatMRPT, an LLM-assisted ITN allocation planning tool based on user requirements, and analyzed post-interaction feedback, examined user digital competencies and workplace resources and identified institutionalization pathways for LLM-assisted intervention planning.

**Methods:** A two-phase mixed-methods study began with software requirements gathering workshops (using a prototype) involving representatives from the National Malaria Elimination Programme (NMEP), State Malaria Elimination Programmes (SMEPs), and implementing partners. Phase two evaluated ChatMRPT through surveys, guided exercises, and focus group discussions with 34 SMEP officers from 28 Nigerian states. Quantitative data were analyzed using descriptive statistics and profile-based comparisons, while qualitative data were analyzed using reflexive thematic analysis to synthesize user experiences of ChatMRPT and identify institutionalization pathways.

**Findings:** Fifty-eight percent (19/33) of participants demonstrated both higher digital competency and adequate workplace resources; the remainder exhibited limitations in one or both domains ([4/33] higher competency/constrained resources; [6/33] higher resources/lower competency). Participants with higher digital competency but constrained workplace resources reported user experiences comparable to those with higher competency and adequate resources, whereas workplace resources alone did not appear to compensate for lower digital competency. Key software requirements included contextual guidance for malaria risk interpretation, operational decision support, and embedded analytical support. Following iterative incorporation of these requirements, ChatMRPT was positively evaluated across participant profiles. Participants viewed institutionalization as dependent on integration into routine malaria planning and adaptability to evolving programme priorities.

**Interpretation:** Many malaria programme officers may already have the foundational competency for LLM-assisted decision support. However, there is room to further strengthen digital competencies while facilitating access to basic workplace resources such as stable internet. Institutionalization of LLM tools may depend on addressing these capacity and infrastructural constraints alongside designing explainable, integrated, and flexible systems. Future research should evaluate long-term integration, sustainability, and effectiveness in routine malaria planning.

**Funding:** This work was funded by the Bill and Melinda Gates Foundation (INV-036449) and the Center for Health Outcomes and Informatics Research (CHOIR), Loyola University Chicago. The funders had no role in the study design, data analysis, interpretation of findings, or preparation of the manuscript.

**Research in context:** *Evidence before this study:* We searched PubMed and Google Scholar for studies published from 2022 onwards using combinations of terms related to LLMs, decision support, malaria planning, implementation, and insecticide-treated nets. Previous studies have integrated epidemiological, environmental, socioeconomic, and operational data to support malaria risk mapping, intervention targeting, and resource allocation, including under resource constraints.

*Added value of this study:* We make three contributions to the evidence based on the use of LLM-assisted tools for malaria intervention planning. First, we describe variation in digital competency and workplace resources and how these relate to user experiences with LLM-assisted tools. Second, we elucidate software design requirements for enhancement of interpretability, contextual exploration, workflow integration, and operational decision support. Third, we identify organizational, technical, and governance conditions shaping institutionalization, including interoperability, leadership support, workflow integration, and adaptability across implementation contexts. Together, these findings inform the design and integration of LLM-assisted decision-support tools for routine malaria programme planning.

*Implications of all the available evidence:* Successful implementation of LLM-assisted risk-informed ITN planning requires stakeholders to interpret and apply analytical outputs within routine planning systems. While previous studies have focused on predictive modelling, optimization, and risk mapping, our findings highlight interpretive support, workflow integration, and stakeholder interaction in translating analytical outputs into planning decisions. Institutionalization further requires stakeholder-centred design, interoperability, organizational support, and capacity strengthening, building on foundational capacity already present within many malaria programmes.

## Introduction

Malaria remains a major public health burden globally, with Nigeria accounting for 24·3% and 30·3% of malaria cases and deaths worldwide (1,2). Although considerable progress has been made in reducing malaria transmission over recent decades (3,4), malaria continues to place significant strain on health systems, livelihoods, and population health outcomes (5,6). Insecticide-treated nets (ITNs) remain one of the most effective and widely implemented malaria prevention interventions across many endemic countries (7–9). ITNs in Nigeria are primarily distributed through periodic mass campaigns coordinated by the National Malaria Elimination Programme (NMEP) and implemented by State Malaria Elimination Programmes (SMEPs) in partnership with implementing partners (9–11).

In Nigeria, increasing financial constraints, competing public health priorities, and heterogeneous malaria transmission have made blanket ITN allocation increasingly unsustainable (1,12,13). When universal coverage is unachievable, the NMEP must determine how limited ITNs should be allocated. Population size remains central to current allocation approaches, but it does not explicitly account for spatial heterogeneity in malaria risk, potentially limiting the prioritization of areas with the greatest epidemiological need (14,15). Differences in how national malaria policies and allocation decisions are translated into practice across states and local government areas due to the involvement of multiple implementing partners, decentralized planning and service delivery structures, and infrastructural constraints further compounded operationalization of ITN prioritization strategies (16,17).

A key barrier for NMEP ownership of the analytical process was the lack of tools to facilitate data aggregation, analysis, interpretation, contextualization, and translation into operational plans for use during mass campaigns (18). This resulted in the development of the Malaria Reprioritization Tool (MRPT) to fill that gap at the national level. However, malaria planning and ITN reprioritization operate within a decentralized programme structure, with SMEPs sharing responsibility for state-level decisions, creating a need to support consistent interpretation and application of MRPT analyses across programme teams with varying analytical experience.

To address this need, we augmented the existing MRPT framework (18) with an LLM-enabled decision-support interface. Together, these components form ChatMRPT (19), combining the MRPT engine, which generates malaria-risk analyses and ITN allocation recommendations, with an LLM interface that enables users to interrogate, interpret, contextualize, and apply these outputs during routine planning. This study examined the development and implementation of ChatMRPT through four aims: (1) characterize user digital competency and workplace resource profiles relevant to LLM-assisted malaria planning; (2) examine variation in perceptions of ChatMRPT across these profiles; (3) synthesize user experiences and stakeholder-informed requirements for LLM-assisted decision-support tools; and (4) identify factors influencing their institutionalization within routine malaria intervention planning.

## Methods

### Overview

MRPT facilitates analysis of deidentified clinical malaria data and visualization and classification of urban settlements by level of formality, enabling integration and mapping of epidemiological, environmental, demographic, and settlement-related indicators. Through an interactive workflow, users generate maps of composite risk scores to identify populations and locations at higher malaria risk and support more targeted, data-informed ITN allocation. ChatMRPT operationalizes the MRPT analytical framework within a Python-based architecture that orchestrates MRPT functions with OpenAI-enabled tools to provide conversational navigation, interpretation of analytical outputs, and stepwise decision support. Analytical outputs, including calculated risk scores and rankings, are generated by the underlying MRPT functions and supplied to the language model for natural-language interpretation and not calculated by the language model. This study evaluated stakeholder interaction with and requirements for LLM-assisted decision support rather than the analytical validity of ChatMRPT-generated outputs. User perceptions should therefore not be interpreted as evidence of analytical accuracy or decision quality, which require separate evaluation.

The study used a two-phase mixed-methods design (Figure 1 and Figure *A1*). Focus group discussions (FGDs) and surveys were conducted across both phases. Phase 1, conducted between February 2024 and early 2025, identified stakeholder-informed requirements for the development of ChatMRPT through engagement with representatives from the National Malaria Elimination Programme (NMEP), State Malaria Elimination Programmes (SMEPs), and implementing partners involved in ITN campaign planning, data management, and implementation. Phase 2, conducted between October 2025 and February 2026 with 34 participants across 28 states including the Federal Capital Territory (FCT) (*Figure A4*), examined SMEP officers’ experiences using ChatMRPT and stakeholder-informed requirements for integrating LLM-assisted decision support into routine malaria planning, particularly ITN mass campaigns. Participants were purposively selected to represent states implementing ITN campaigns under the Global Fund (GF), Immunization Plus and Malaria Progress by Accelerating Coverage and Transforming Services (IMPACT), and the President’s Malaria Initiative (PMI), thereby capturing different campaign funding arrangements and distribution approaches. Although six of the 34 participants had also participated in Phase 1, Phase 2 was not designed as a longitudinal follow-up.

**Figure 1:**
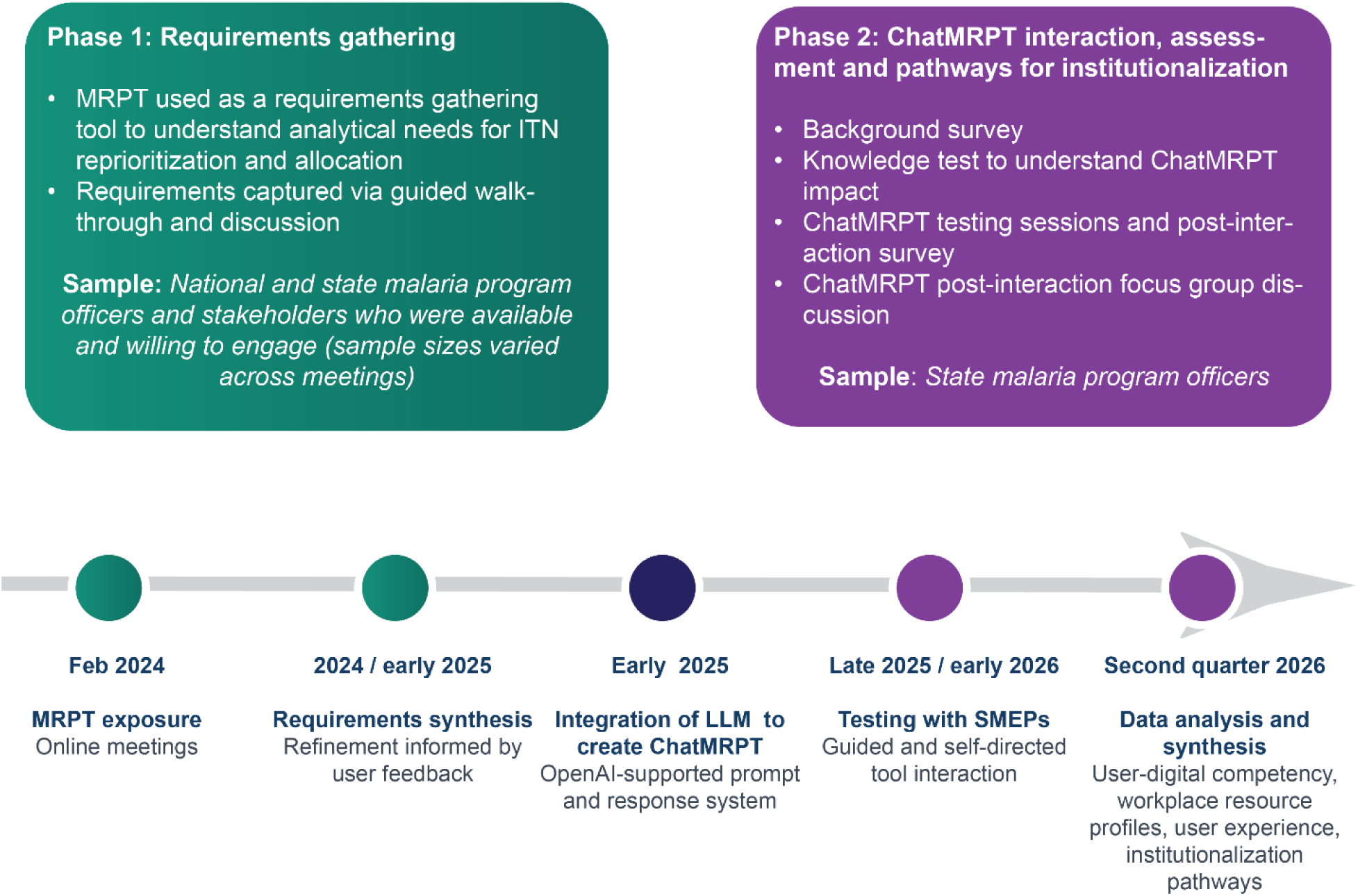
Study concept and timeline illustrating requirements gathering and iterative development, refinement, and evaluation process of Chat-Malaria Reprioritization Tool.

### Data Collection

Data were collected through audio recordings of guided tool walkthroughs and focus group discussions (FGDs), and responses to online surveys.

In Phase one, stakeholders from national and state levels and selected implementing partners engaged in a series of virtual guided walkthroughs of MRPT to enable feedback gathering for further development of an analytical tool to facilitate ITN reprioritization and allocation during mass campaigns. These activities also served to identify system requirements, workflow constraints, and decision-making challenges within existing ITN distribution processes, particularly in urban areas, which was the focus of ITN reprioritization. The insights generated informed modifications to the tool, including the addition of an LLM-powered component, resulting in the development of ChatMRPT.

In Phase two, the study team conducted virtual FGDs and ChatMRPT testing sessions across multiple rounds with 34 SMEP officers. One participant did not complete all survey components, and the missing data were assumed to be missing at random (MAR).

To characterize factors likely to influence routine use of ChatMRPT in malaria planning and ITN reprioritization, we developed a study-specific descriptive profiling framework. Given the lack of frameworks and tools specific to our study, we drew on existing technology acceptance, organizational readiness, and implementation frameworks, which provide important insights into technology adoption and implementation (20), and emerging AI readiness research that highlights the importance of both digital skills and enabling infrastructure for AI adoption (21). The framework comprised two complementary dimensions: (i) user digital competency, reflecting participants’ digital skills, experience with analytical and geospatial tools, and routine data use; and (ii) workplace resource availability, reflecting access to the minimum infrastructure required for routine platform use. The selected indicators were informed by the technical requirements of ChatMRPT and stakeholder input obtained during the co-design workshops. Following the initial FGDs, participants completed a survey (*Appendix Three, Instrument Two*) to collect information on these two dimensions. The resulting data were used to construct descriptive participant profiles for subsequent analyses.

Participants then engaged with ChatMRPT through a two-part workflow comprising a guided session and an independent-use session. During the guided session, facilitators demonstrated the complete ChatMRPT workflow (*Appendix Two*) and completed another survey to gauge their perspectives on ChatMRPT (*Appendix Three, Instrument Two*).

FGDs explored participants’ experiences using ChatMRPT and the conditions required for its institutionalization into routine malaria planning (*Appendix Three, instrument Four*).

### Data Analysis

#### Phase 1

Recordings were transcribed verbatim and reviewed to identify recurring concepts, operational challenges, and user requirements. These were synthesized into requirement themes to identify actionable development priorities that informed the creation of ChatMRPT. Key findings are reported and illustrated through representative quotes.

#### Phase 2

ChatMRPT post-interaction FGDs were analyzed using inductive thematic analysis. Initial codes were developed from recurring concepts identified during transcript review and organized into a preliminary codebook. Two independent (LM and EAB) coders applied the codebook, resolving discrepancies through discussion and iterative refinement of thematic definitions over three coding rounds. Inter-coder reliability was assessed using Cohen’s kappa, improving successive refinements of the coding framework. The final coded dataset demonstrated moderate agreement (κ = 0·604; n = 437 coded text segments). Coding, aggregation, and thematic synthesis were conducted using spreadsheet-based coding matrices and custom Python workflows. Phase 2 findings were presented alongside those from Phase 1.

#### User digital competency and workplace resource profile analysis

Two descriptive participant profiles were constructed: a user digital competency profile and a workplace resource profile. The user digital competency profile characterized participants’ capacity to engage with an LLM-assisted decision-support tool using three self-reported characteristics: (i) computer skills, (ii) frequency of working with data, and (iii) previous use of analytical, geospatial, and digital decision-support tools. Unique combinations of these characteristics were grouped into nine ordered digital competency profiles (P1–P9), ranging from lower (P1) to higher (P9) competency. Profiles were ordered hierarchically, prioritizing computer proficiency, followed by frequency of routine data use and previous experience with digital analytical tools. Computer proficiency and routine data use were prioritized because they reflect the ability to navigate the platform and critically engage with analytical outputs, whereas platform-specific digital skills can be strengthened through training.

Workplace resource profiles were constructed from four indicators representing infrastructure relevant to routine ChatMRPT use: access to a laptop or desktop computer, smartphone or tablet, stable internet, and stable electricity. Distinct combinations were grouped into six ordered profiles (R1–R6), ranging from lower (R1) to higher (R6) resource availability. Profile ordering considered both the number and functional relevance of available resources, with stable internet and electricity prioritized over access to multiple device types because computer and smartphone/tablet access provide partially overlapping means of accessing the platform. Profiles with at least three resources (R4–R6) were classified as higher-resource profiles.

Both profile sets were used descriptively to characterize variation across participants. A cross-tabulation of digital competency and workplace resource profiles was then used to examine their joint distribution, with cell values expressed as the proportion of all study participants. To support interpretation, profile codes were ordered from lower to higher levels of digital competency and workplace resource availability.

#### Analysis of Likert-scale responses from ChatMRPT post-interaction surveys

Likert-scale responses reflected participants’ immediate perceptions following interaction with ChatMRPT and were interpreted as initial impressions rather than assessments of sustained use. Survey questions were grouped into five conceptually related evaluation domains, with negatively worded items reverse-coded so that higher scores represented more favorable perceptions. Responses within each domain were averaged per participant, with mean scores ≥4 classified as positive, and summarized overall and by digital competency, workplace resource, and joint competency-resource profiles. Higher digital competency (P6–P9) and workplace resource (R4–R6) profiles represented at least intermediate computer skills, frequent routine data use, and access to three or more workplace resources. Supplementary analyses examined positive responses and Likert-scale distributions of individual survey items underlying these summaries. Fisher’s exact tests examined associations between positive perceptions and digital competency and workplace resource profiles within each domain, with Benjamini–Hochberg adjustment for multiple comparisons.

#### Pathways and barriers to institutionalization

We analyzed qualitative responses on broader system-level considerations for the institutionalization of ChatMRPT. Data coded within this theme were subsequently re-examined to identify organizational, technical, and operational subthemes describing requirements for institutionalization. Representative quotations were selected to illustrate each domain.

### Funding

The study was funded by the Bill and Melinda Gates Foundation (Investment ID: INV-036449) and the Center for Health Outcomes and Informatics Research (CHOIR). The funders had no role in the design or analysis of this study

## Results

Results are presented according to the study objectives rather than by data collection sequence. We first describe participant digital competency and workplace profiles, followed by a joint presentation of stakeholder-derived design requirements and user perceptions of ChatMRPT post-implementation of these design requirements. We conclude with conditions for institutionalization identified during stakeholder discussions.

### User profile: digital competency and workplace resource availability

The 34 phase 2 participants were predominantly engaged in monitoring and evaluation–related roles (30/34 [88·2%]), reflecting a technically oriented user group; 47·1% held a Master of Science degree and 38·2% a Bachelor of Science degree (*Appendix, Figure A5*). Most respondents (88%) also reported frequent involvement in ITN planning and distribution activities (often or very often; Figure 2A). Participants had prior experience with a range of digital tools, with analytics platforms most commonly reported (25%) and prior use of AI tools uncommon (4%; Figure 2B).

**Figure 2:**
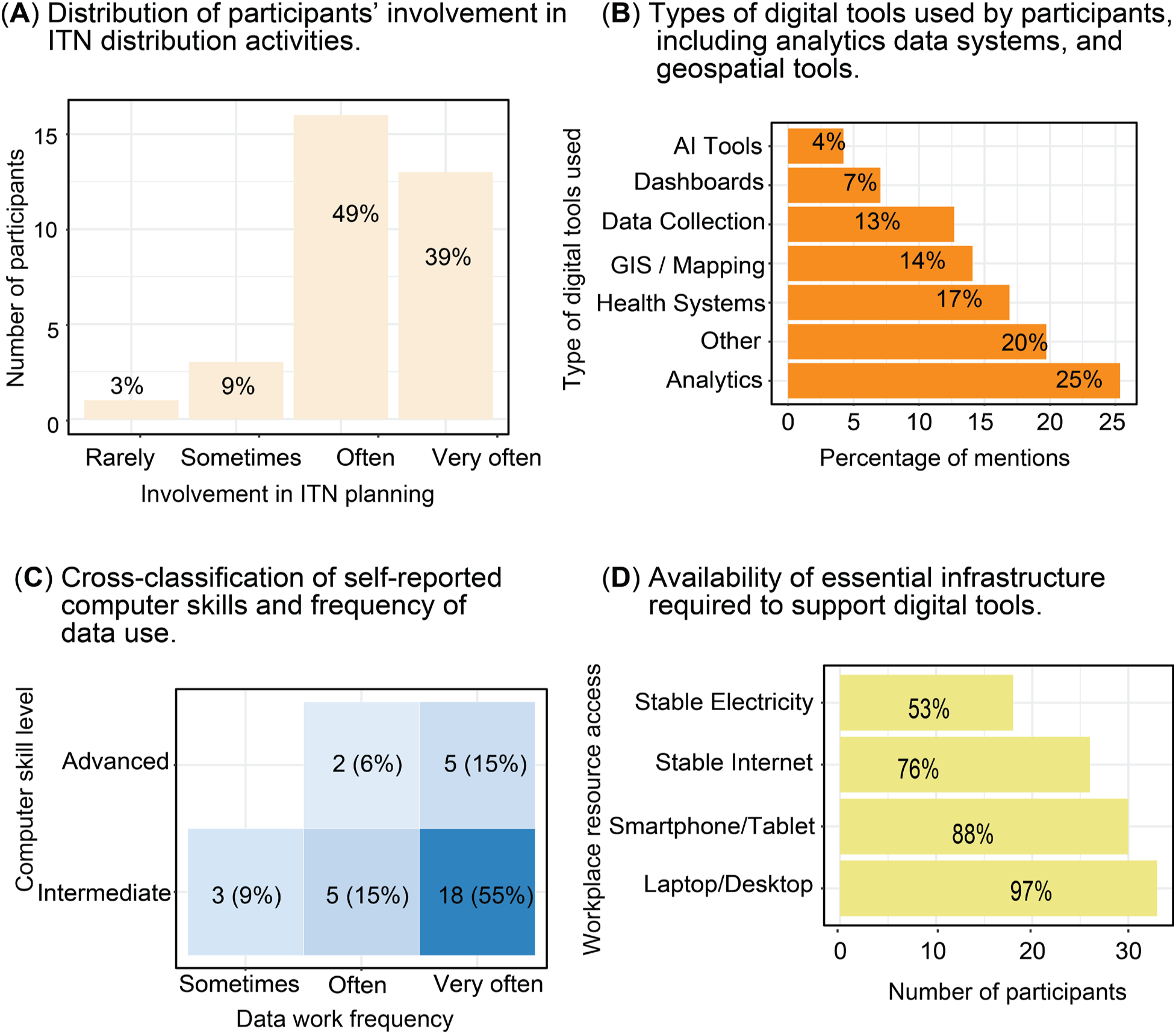
Profiling users’ role in ITN distribution, digital tool and data use, and workplace resource access. (A) Levels of involvement in ITN planning (B) Reported use of digital tools (C) Matrix combining self-reported computer skill level and frequency of data use. (D) Access to infrastructure. * Health systems is short for Health information systems such as DHIS2. See Appendix for details on each category.

Figure 2C shows substantial variability in self-reported computer skills and frequency of data use across participants. Most participants reported intermediate computer skills combined with very frequent data use (54·5%), while smaller proportions reported advanced skills or less frequent data use. Although routine data use was common, participants differed in their self-reported computer skill levels.

Access to digital infrastructure was generally high but uneven across participants (Figure 2D). While nearly all participants reported access to laptops/desktops and most had smartphones/tablets, access to stable internet connectivity and electricity was less consistent. Although most participants reported access to devices and internet connectivity (Figure 2D), qualitative findings and supplementary analyses (Figure A6) suggest that the quality of internet connectivity, rather than access alone, may be an important constraint in some work settings.

Figure 3A shows variation in workplace resource profiles, with 45% (15/33) of participants classified in the highest workplace resource profile (R6). Figure 3B summarizes participants’ digital competency profiles, derived from self-reported computer skills, frequency of data use, and previous use of digital decision-support tools. Overall, 69·7% (23/33) of participants were classified within the higher digital competency profiles (P6–P9), while the remainder fell within profiles P1–P5.

**Figure 3:**
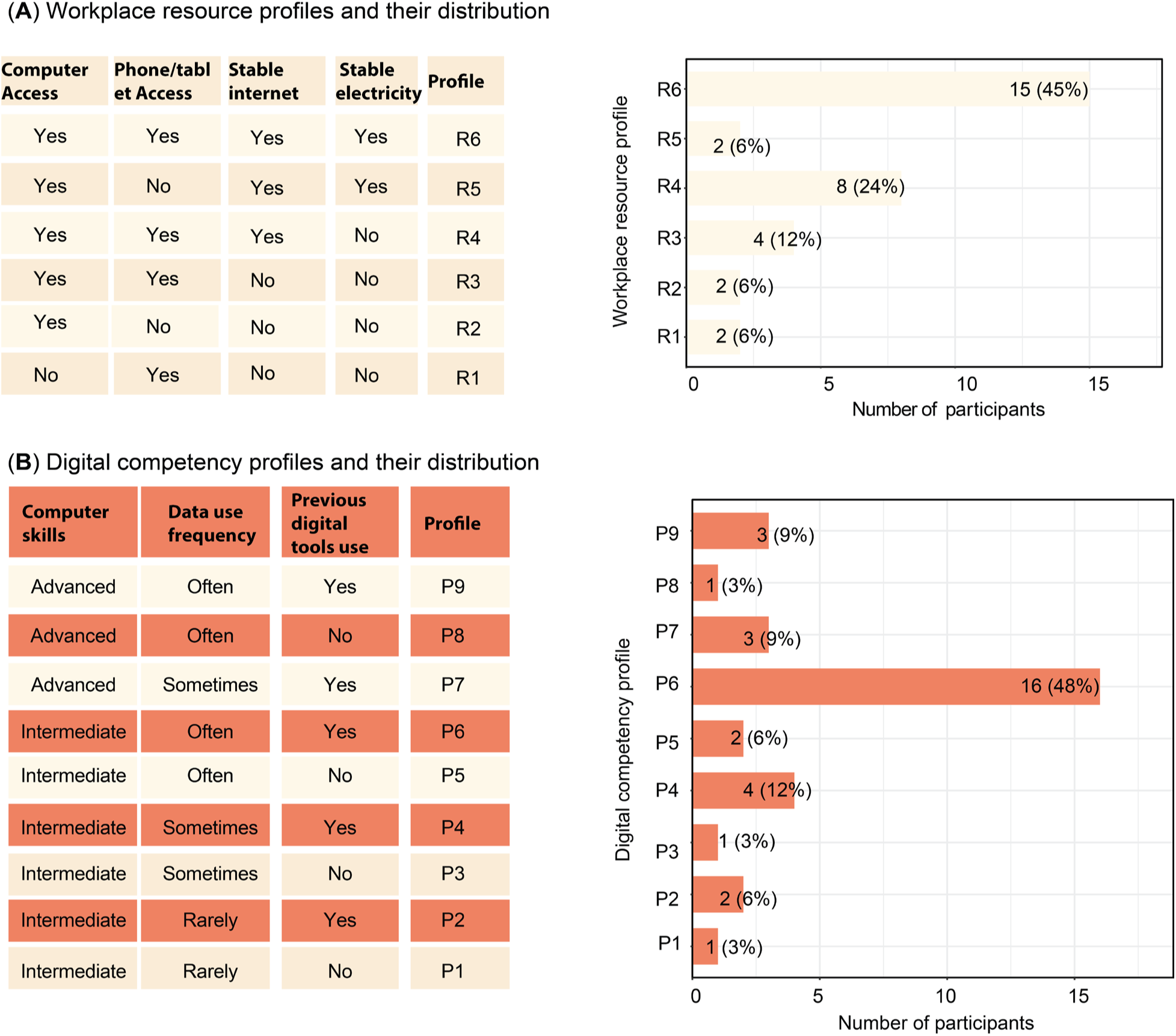
Participants’ digital competency and workplace resource characteristics. (A) Workplace resource profiles derived from self-reported access to computers, mobile devices, stable internet, and electricity. (B) Digital competency profiles derived from self-reported computer skills, frequency of data use, and previous use of digital decision-support tools.

When digital competency and workplace resource availability were considered jointly (Figure 4A), participant competency and resource profiles were heterogeneous. Figure 4B summarizes the four profile categories derived from the joint distribution. Overall, 57·6% of participants were classified within the upper-left quadrant corresponding to digital competency profiles P6– P9 and workplace resource profiles R4–R6. Among the remaining 42·4%, three distinct profiles emerged: 18·2% had adequate workplace resources but lower digital competency, 12·1% had higher digital competency but constrained workplace resources, and 12·1% had limitations in both dimensions.

**Figure 4:**
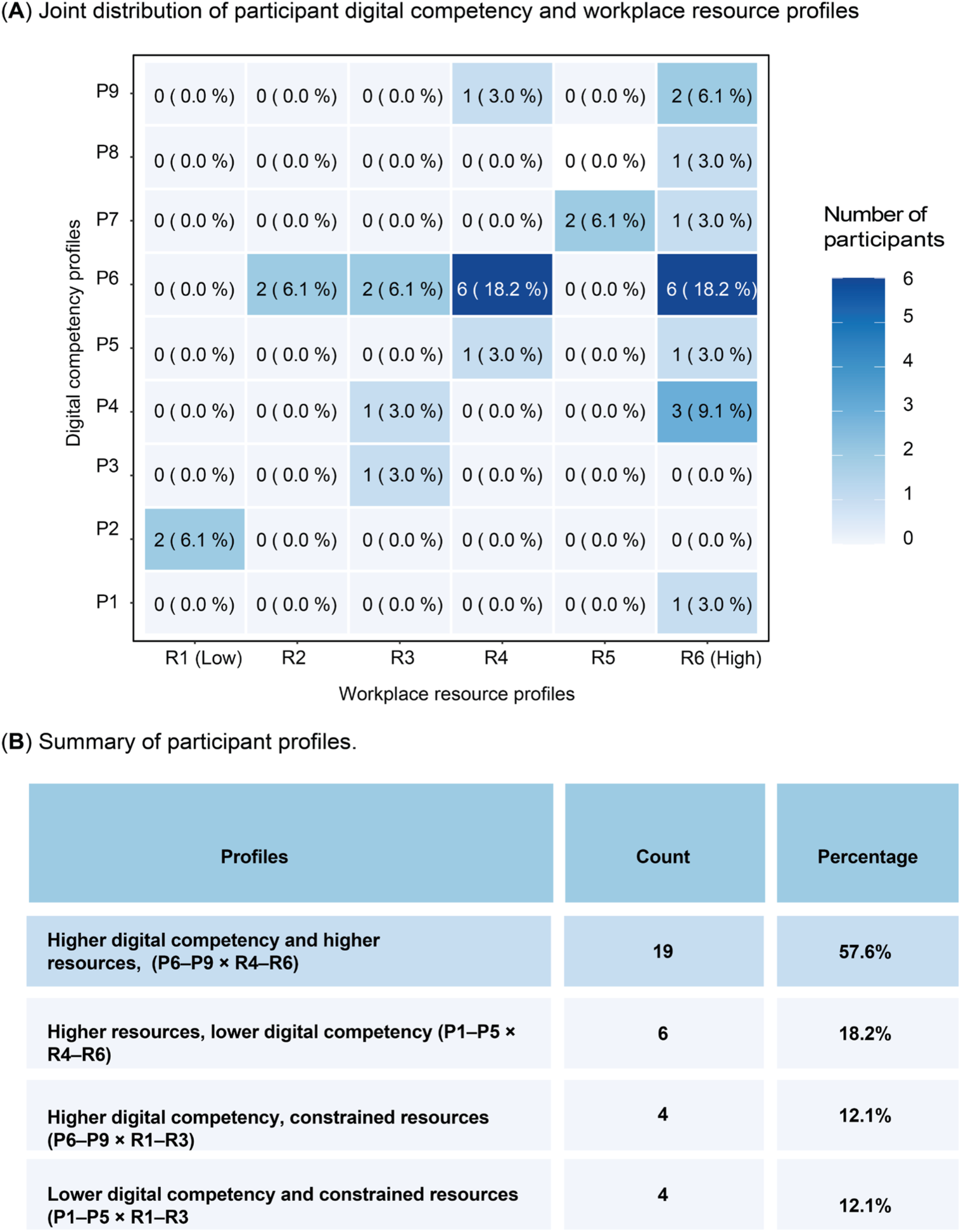
Joint distribution of participant digital competency and workplace resource profiles. (A) Heatmap showing the joint distribution of digital competency profiles (P1–P9) and workplace resource profiles (R1–R6). Cell values represent the number (percentage) of participants (n = 33) within each profile combination. Profiles are ordered from lower to higher digital competency and workplace resource availability. (B) Summary of participant profiles derived from the joint distribution of digital competency and workplace resource availability, showing the proportion of participants with higher digital competency and workplace resource profiles and the three remaining profile combinations.

### Stakeholder-informed development and feedback following interaction with ChatMRPT

#### User perception of ChatMRPT by digital competency and workplace resources

Participants’ perceptions of ChatMRPT were assessed among the participants (Figure 5). At the participant level, positive perceptions were highest for ease of navigation (93%) and willingness to use and recommend the platform (86%), followed by ease of understanding and interaction (79%) and system complexity and support (62%). Ease of learning and confidence was less consistently positive (48%; Figure 5A). Item-level analyses provided additional context for these domain-level summaries: most participants reported confidence in using ChatMRPT, while responses were more varied regarding the amount they needed to learn before using the platform effectively (Figure A8).

**Figure 5.**
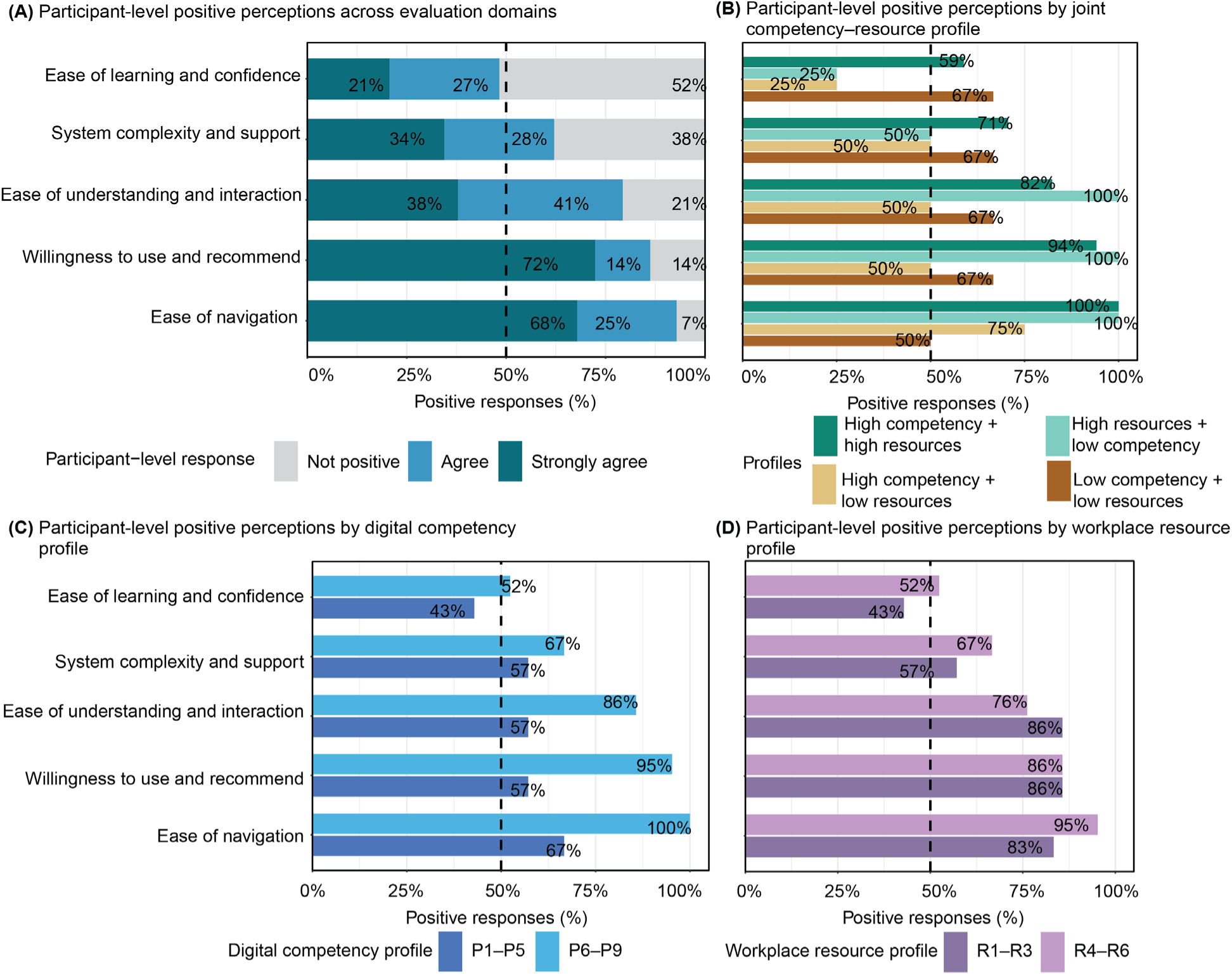
Participant-level perceptions of ChatMRPT across evaluation domains and participant profile groups. (A) Positive perceptions across the five evaluation domains, based on participants’ mean responses within each domain; mean scores ≥4 were classified as positive. The dashed line indicates 50% of participants. (B) Positive perceptions stratified by the four digital competency–workplace resource profile combinations (* Joint-profile estimates are descriptive and should be interpreted cautiously because of small cell sizes.). (C) Positive perceptions stratified by digital competency profile (P1–P5 vs P6–P9; n=10 vs 23). (D) Positive perceptions stratified by workplace resource profile (R1–R3 vs R4–R6; n=8 vs 25).

Positive perceptions generally increased with digital competency, with the largest differences between higher- and lower-competency profiles observed for willingness to use and recommend ChatMRPT (95% vs 57%), ease of understanding and interaction (86% vs 57%), and ease of navigation (100% vs 67%; Figure 5C). Differences by workplace resource profile were smaller and inconsistent in direction; perceptions of willingness to use and recommend were identical across resource groups (86%), while understanding and interaction was higher among participants with fewer workplace resources (86% vs 76%; Figure 5D). Joint competency– resource profiles similarly showed that positive perceptions were not restricted to participants with both higher competency and higher resources, although estimates varied considerably across the smaller profile groups (Figure 5B).

Exploratory Fisher’s exact tests suggested unadjusted differences by digital competency profile for ease of understanding and interaction (p=0·011) and ease of navigation (p=0·043); however, neither remained statistically significant after Benjamini–Hochberg adjustment (adjusted p>0·05). No significant differences were observed by workplace resource profile across the evaluation domains (all adjusted p=1·00).

#### Qualitative synthesis of user feedback

User requirement analysis identified three domains: guidance for malaria risk analysis, integration of local expertise with systematic evidence, and support for translating malaria risk analyses into operational decisions. The following sections describe participants’ experiences using ChatMRPT after these requirements were incorporated into the platform.

#### Guidance for conducting malaria risk analyses

Stakeholders initially described substantial challenges understanding how malaria-risk analyses were generated, including uncertainty about required data inputs, variable selection, and the analytical processes used by the MRPT. One participant noted difficulties understanding the required inputs, asking, “*How do we assemble these variables? How do we know which variables we need to upload?*” while another noted, “*I’ve tried it on my own to visualize this data, but how it is being arrived at, I find it very difficult.*”

Following interaction with ChatMRPT, participants reported greater confidence in conducting malaria risk analyses and generating outputs. They described the system as easy to use, highlighting its guided prompts and reduced training requirements. One participant explained that “*it has really helped me so well (…) On my own now I can just put on that data and check where the malaria is at high risk,*” while another noted that “*the guide to the prompt is also good (…) even if you’re a new user, you may not really go through a lot of training.*” Responses were frequently described as “*clear*,” “*helpful*,” and “*easy to understand.”*

Participants also reported greater confidence in communicating and defending analytical outputs. One participant stated that “*ChatMRPT will go a long way to help with the confidence level in being able to explain these to stakeholders,*” while another described the outputs as providing “*good defense (…) evidence based decision[s]*” during microplanning/allocation discussions.

#### Balancing local expertise with systematic evidence-based analysis

The requirement gathering participants emphasized that analytical inputs and (or) outputs needed to reflect local epidemiological and operational realities to be useful for decision-making. During engagements with the MRPT, stakeholders highlighted the importance of contextualizing malaria-risk variables included in the composite scoring and allowing flexibility in how reprioritization decisions were explored across states. One participant noted that *“…there are more water bodies in [State A] compared to the likes of [State B]…,*” while emphasizing that risk factors may operate differently across settings. Others argued that “*the section for parameters [should] be flexible so that, based on the context in the state and agreement with stakeholders, we can decide areas to deprioritize,*” highlighting the need for locally informed decision-making rather than reliance on fixed variables.

While local contextual knowledge was viewed as essential, participants also recognized that it could influence decision-making in ways that may not fully reflect the broader evidence base. As one stakeholder explained, "*I would just take temperature (…) because that’s what I feel drives my transmission"* illustrating how users may gravitate toward variables they know or perceive as important rather than considering a broader range of factors based on the research literature.

SMEPs described ChatMRPT as facilitating interactive exploration of state-specific malaria-risk patterns and intervention-targeting scenarios. Participants reported using the conversational interface to investigate “*what is happening in our state,*” by examining and validating high-risk wards, and generating visualizations that reflected locally derived surveillance data. Rather than relying on predetermined allocation approaches, they described using ChatMRPT to interrogate state-specific epidemiological patterns and support discussions around locally appropriate reprioritization decisions.

This contrasted with the requirements phase, where participants requested greater flexibility to account for differences between states, likely reflecting the NMEP’s needs for cross-state comparisons. During the ChatMRPT evaluation, participants no longer requested additional contextual customization, instead participants focused on how ChatMRPT enabled exploration and interpretation of their own state’s epidemiological patterns.

#### Translating malaria risk analyses into operational decisions

During stakeholder engagements, participants emphasized the need for decision-support tools that translate malaria-risk analyses into actionable planning decisions. One participant noted that tools should “*reflect reality on the ground and direct us to appropriate solutions.*”

Following interaction with ChatMRPT, participants reported that the platform addressed these requirements by supporting intervention targeting, reprioritization, and operational planning. SMEPs described using the tool to interpret risk outputs, translate rankings into prioritization plans, and explore alternative allocation scenarios to inform resource allocation decisions. One participant explained that the tool “*gives you direction where to target your interventions,”* while another described it as “*useful for planning and decision making.*” Another participant described the platform as “ *stating the highest risk ward and giving a guide to net distribution,*” while another noted that it was “*a very useful app that will go a long way helping us to know areas where we should channel our strength and interventions to.*”

Another participant described ChatMRPT as helping address a recurring planning challenge in which “*areas that don’t need net actually have more nets and areas that need more net don’t even have enough nets.*” By linking analytical outputs to operational questions, participants perceived the platform as supporting more transparent and evidence-informed allocation discussions.

### Conditions for institutionalization

Although participants viewed ChatMRPT positively and recognized its value for malaria planning, they emphasized that successful institutionalization depends on more than a technically capable platform. They identified six organizational, technical, and governance conditions grouped into two domains: embedding LLM-assisted decision support within routine malaria planning and sustaining its use across evolving ITN implementation contexts (Figure 6).

**Figure 6:**
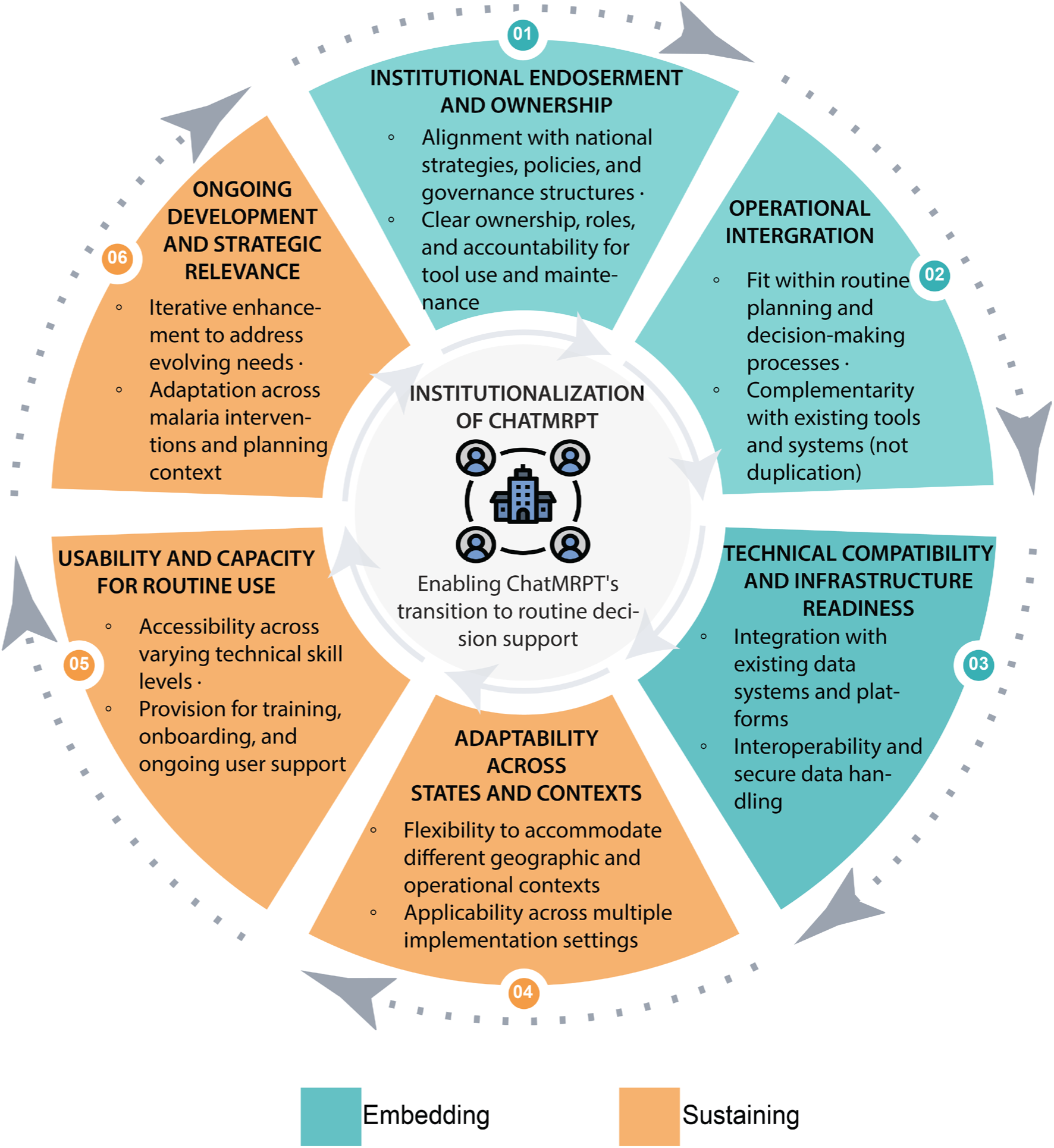
Conditions for Institutionalizing ChatMRPT in Routine Malaria Planning Systems.

#### Embedding LLM-assisted decision support

<u>Institutional endorsement and ownership</u> were a prerequisite for institutionalization. Participants emphasized that support from the national coordinator and state programme manager would be necessary to facilitate coordinated adoption and routine use. As one participant noted, “*after we have buy-in from the national level down to the SMEP level (…) rolling out at the bottom is very easy.*” Participants also observed that implementing partners often use different planning systems, noting that “*we have different partners we are working with in the state, and every partner has their tools they use for different campaigns,*” highlighting the need for alignment across implementing partners, who play a central role in determining which planning tools are adopted across campaigns.

<u>Operational integration</u> within existing planning processes was viewed as equally important. Participants emphasized that ChatMRPT should complement rather than duplicate existing systems, asking, “*are we going to have a parallel tool or are we working in tandem with what WHO provided?”* SMEPs also highlighted the need to engage other personnel directly involved in campaign planning and implementation, noting that “*another set of persons (…) that need to understand this tool is the logistics officer (…) because they are the ones that do the major planning of this campaign*.”

<u>Technical compatibility and infrastructure availability</u> were identified as foundational requirements for institutionalization. Participants emphasized the importance of interoperability with existing data systems and workflows, noting that the platform should “*integrate with the existing health information infrastructure*” and be “*linked to other tools like DHIS2.”* Several stakeholders highlighted practical considerations related to data management, including compatibility with existing data formats, streamlined data uploads, and the ability to retain uploaded datasets across analytical sessions. Infrastructure was frequently discussed, with participants noting that performance depended on internet connectivity: “*if your internet is stable, it works well… once your internet is slow, it has a longer response time.”* Others emphasized the need for compatibility across devices and platforms, including browser- and mobile-based access, to support use across diverse operational settings.

#### Sustaining use across contexts and evolving programmatic needs

<u>Adaptability across states and implementation contexts</u> was also a recurring theme. Participants noted that operational needs vary across settings, with one participant observing that “*it’s not every state (…) some states are having challenges, especially when it comes to net distribution.*” Stakeholders emphasized the importance of ensuring that the platform could support different programmatic needs across these settings.

<u>Usability and capacity for routine use</u>, encompassing simplified interpretation, continued capacity strengthening, and trust, were identified as key enabling factors for sustained engagement. Participants valued the embedded prompts and guidance, describing them as features that supported navigation and reduced the effort required to interact with the platform. With one participant specifically saying, “*… notice the prompts (…) the prompt is good…*” Participants emphasized the importance of continued training and hands-on experience to build confidence and proficiency, noting that “*continuous training and practice (…) is one of the most useful ways so that one can master it.*” Stakeholders also highlighted the need to extend training beyond SMEP staff to include program managers, funders, and other decision-makers involved in malaria planning. Trust in the platform was considered equally important, with participants emphasizing the need for “*a platform that people have trust in*” and confidence that the outputs are reliable, credible, and appropriate for informing operational decisions.

<u>Ongoing development and strategic relevance</u> were also important determinants of long-term institutionalization. Participants emphasized that sustained use would depend on the platform evolving alongside changing programme priorities and user needs. While ChatMRPT was developed to support ITN reprioritization, stakeholders envisioned broader applications across malaria control, with one participant suggesting, *“let’s integrate it for SMC, ITN and other routine activities,”* and another noting that *“other modules that relate to malaria programs can also be incorporated.”* Participants also emphasized that continued refinement and expansion of functionality would strengthen long-term institutionalization, with one stakeholder remarking that *“if we can get these additional features (…) then it can be adopted 100% as a planning tool.”*

## Discussion

This study characterized user digital competency and workplace resource profiles, identified stakeholder-informed design requirements, synthesized user perceptions of ChatMRPT and examined conditions for institutionalizing LLM-assisted malaria planning.

### User competency and workplace resources: a differentiated picture

Participants reported little resistance to LLM-assisted decision-support tools or digital systems as a major barrier. Instead, while previous studies have emphasized digital literacy as a key barrier to digital health adoption (22–24), sustainable institutionalization of tools such as ChatMRPT may depend more on addressing contextual constraints, including infrastructure, connectivity, and ongoing capacity strengthening.

Among participants outside the higher digital competency–higher workplace resource profile, barriers to LLM-assisted decision-support tools planning arose from different combinations of individual digital competency and workplace resources rather than a single constraint. Consistent with broader digital health implementation research, which identifies workforce capacity and organizational context as important determinants of technology adoption (22–24), some users may benefit primarily from capacity strengthening, whereas others require workplace infrastructure improvements before engaging effectively with LLM-assisted tools. These findings suggest that implementation strategies should be tailored to participant profiles rather than adopting a uniform deployment model. Positive perceptions across all digital competency groups further indicate that lower digital competency alone should not preclude routine use when appropriate support is available.

### Design considerations: what requirements analysis reveals

Effective LLM-assisted malaria planning tools require both accurate analytical outputs and transparent, explainable processes that enable users to understand how recommendations are generated. This complements existing malaria decision-support literature, which has concentrated primarily on predictive accuracy and optimization (25) with comparatively less attention paid to whether and how analytical outputs are made transparent, explainable, and translatable into the operational realities of routine programme planning. A similar challenge has been identified in molecular and genomic surveillance, where even well-developed data streams remain underused in malaria programme decision-making when they are not translated into actionable guidance (26).

Within our sample, participants treated transparency and explainability as a necessary complement to accurate outputs rather than a secondary consideration, supporting an argument that explainability functions as a core design requirement rather than an optional usability feature. In ChatMRPT’s case, this took the specific form of natural-language explanation, guided prompts, and contextual exploration, plain-language re-explanation of outputs rather than formal interpretability methods such as feature attribution (27). Advancing MRPT into ChatMRPT shifted the tool’s role from generating malaria-risk outputs to supporting their interpretation and application, which participants described as reducing the cognitive effort required to translate analytical outputs into operational decisions.

This distinction was also evident in the quantitative findings. Although most participants reported feeling confident using ChatMRPT, responses were more varied regarding whether they needed to learn many things before using it effectively, resulting in lower participant-level positive perceptions for the combined ease-of-learning and confidence domain. This suggests that confidence in interacting with an LLM-assisted platform may coexist with a perceived need for additional learning to interpret and apply its outputs effectively. Together with participants’ qualitative emphasis on guided prompts, contextual explanations, and plain-language interpretation, this pattern suggests that the relevant implementation challenge may extend beyond basic platform usability to ensuring sufficient analytical and contextual support for translating outputs into programme decisions. Explainability may therefore represent a core requirement for institutionalizing LLM-assisted decision support rather than an optional usability feature, consistent with implementation science literature emphasizing compatibility with existing workflows (27–29).

A further pattern in participants’ accounts was that ChatMRPT improved their confidence in communicating and defending analytical outputs to others. Because ITN allocation decisions involve negotiation among implementing partners and political stakeholders, this suggests LLM-assisted decision-support tools may provide value not only by supporting internal operational decision-making but also by strengthening SMEP staff’s capacity for evidence-informed\\\\\\ advocacy in cross-partner settings, a function distinct from, and additional to, analytical support itself.

### Institutionalization: systems, not users, constrain adoption

SMEPs identified six stakeholder-derived conditions for institutionalizing LLM-assisted decision-support tools, organized into embedding and sustaining domains. These findings extend the existing digital health literature on technology adoption in LMIC settings (22–24) by providing empirically grounded evidence on implementation requirements for institutionalizing LLM-assisted decision-support tools within routine malaria programme planning.

The embedding domain highlights that institutionalization depends on governance alignment, operational integration, and technical compatibility, not user capability alone. Leadership endorsement from the National Coordinator and State Programme Coordinators, together with complementarity rather than duplication of existing planning systems, was viewed as a prerequisite for successful institutionalization. These findings align with implementation science frameworks emphasizing that successful technology integration depends on organizational alignment and compatibility with existing workflows (27–29) while illustrating how these principles operate within the governance structures of Nigeria’s multi-partner malaria planning environment.

The fragmented implementation ecosystem that characterizes malaria planning in Nigeria (30) (multiple funders, implementing partners, reporting systems, and operational mandates) creates challenges for institutionalization that are not fully addressed in the existing literature. Existing digital health research acknowledges that fragmented ecosystems constrain adoption (24,31) but rarely examines how this fragmentation shapes the specific conditions under which tools, particularly LLM-assisted decision-support tools can be embedded within routine planning processes. This fragmentation is not a system weakness. Rather, it reflects the distribution of responsibilities across multiple funders and implementing partners, allowing the financial and operational demands of malaria planning to be shared. However, it requires LLM-assisted decision-support tools to operate within this existing ecosystem rather than creating parallel structures.

The sustaining domain emphasizes adaptability, ongoing capacity development, and continued strategic relevance. Long-term institutionalization depends on the platform evolving alongside programme priorities while maintaining reliable and trustworthy decision support.

### Human oversight remains essential

Participants emphasized that trust in LLM-generated outputs depended on their ability to be interpreted, validated, and reconciled with local programme knowledge. This aligns with broader literature cautioning against the uncritical use of LLMs in public health decision-making (32–35). These findings suggest that human oversight should be viewed as a core component of LLM-assisted decision support, with users critically evaluating outputs rather than accepting them uncritically (36).

### Limitations and future work

ChatMRPT was evaluated through structured demonstrations, onboarding, and guided interaction rather than routine operational use. Consequently, user experiences may differ during sustained implementation, particularly where training and ongoing support vary. Longitudinal evaluations embedded within routine malaria planning are therefore needed to assess independent use, workflow integration, users’ ability to validate and contextualize LLM-generated outputs, and longer-term institutionalization. Although analytical outputs were generated by predefined MRPT functions rather than the language model, this study did not systematically evaluate the fidelity with which the LLM communicated and interpreted these outputs. Future evaluations should assess numerical fidelity and consistency of LLM-generated interpretations before routine implementation.

The quantitative findings should also be interpreted in light of the study design. Digital competency, workplace resources, and user perceptions were self-reported and may not fully reflect participants’ capabilities or experiences during independent use. In addition, survey questions were grouped into conceptually related evaluation domains but were not designed or validated as multi-item scales. Participant-level averages should therefore be interpreted as descriptive summaries rather than validated measures of underlying constructs, with item-level analyses provided to retain the response patterns underlying these summaries. Estimates from the joint competency–resource profiles should similarly be interpreted cautiously given the small numbers in some profile groups.

Finally, although coding reliability was strengthened through independent coding and consensus procedures, thematic analysis remains inherently interpretive. Participants represented diverse roles within Nigeria’s malaria programme, but findings may not generalize to other implementation settings. Analyses were also not stratified by campaign funder to protect confidentiality, limiting assessment of how implementation context influenced digital competency, workplace resources, and perceptions of ChatMRPT.

### Conclusion

Although digital competency and workplace resource profiles varied, SMEPs across participant profiles perceived ChatMRPT positively, suggesting that conversational LLM-assisted decision-support tools could support interpretation of malaria-risk analyses and evidence-informed planning. Sustainable institutionalization, however, extends beyond user capability. Stakeholder-derived requirements – including usability, workflow integration, interoperability, organizational support, and continued capacity strengthening – highlight the importance of designing LLM-assisted decision-support tools to operate within existing programme systems and evolve alongside programme priorities.

## Data Availability

The quantitative survey data, analysis scripts, and associated metadata supporting this study are publicly archived on Zenodo (https://doi.org/10.5281/zenodo.21895518). Additional study data are available from the corresponding authors on reasonable requests. TPR data are subject to data ownership restrictions, and requests for access should be directed to the National Malaria Elimination Programme.

## Declarations

### Ethics approval and consent to participate

Ethical approval for the survey was granted by the Institutional Review Board at Loyola University Chicago Health Sciences Division (Approval Number: 291901 and 218561) and authorization on transcription usage was obtained retrospectively from the Institutional Review Board at Loyola University Chicago Health Sciences Division (Approval Number: 219385) to cover transcribed audio recordings for phase one. The study conformed to the ethical principles of the Helsinki Declaration for the protection of participants. Participants were purposively identified by the National Malaria Elimination Programme (NMEP) based on their roles in malaria programme planning and implementation. Eligible individuals were invited to participate and provided study information before the sessions by the Urban Malaria Team. In accordance with the approved study procedures, informed consent was implied through participants’ voluntary agreement to participate and attend the study sessions.

## Consent for publication

Not applicable

## Acknowledgements

The authors sincerely thank the National Malaria Elimination Programme (NMEP), representatives from the Federal Ministry of Health (FMoH), State Ministries of Health (SMoH), State Malaria Elimination Programmes (SMEPs), and other policymakers who supported this work through participant recruitment, training activities, participating, and prototype evaluations. We are especially grateful to all study participants for their valuable insights, feedback, and engagement, which were instrumental in the refinement and development of MRPT and consequently ChatMRPT. We also acknowledge the contributions of members of the Urban Malaria Project, whose expertise and input supported the design and development of these tools. Support for personnel effort contributing to this work was provided in part through funding from the Center for Health Outcomes and Informatics Research (CHOIR).

## Authors contributions

LM conceived the study, co-developed the study design and data collection tools, developed the initial prototype, conducted the analyses, interpreted the results, and produced the first draft of the manuscript. BOB advanced the prototype into ChatMRPT, developed and maintained the platform, and assisted with transcription and curation of focus group recordings. HATA, EAB, GE, YJ, and GL facilitated focus group discussions, provided feedback on application development, contributed to thematic coding, and assisted with transcription and curation of recordings. CO and IM supported study implementation and provided operational guidance throughout the study. IDO, as Principal Investigator, supported the conceptualization of the study, co-developed the study design and data collection tools, contributed to the conception and refinement of the prototype, oversaw study implementation, provided scientific and strategic supervision throughout the project, critically reviewed the analysis and interpretation of findings, and provided substantial input into manuscript structure and content. LM and IDO contributed to the interpretation of findings and refinement of the analytical framework. More than one author (LM, IDO, BOB, HATA, and EAB) accessed and verified the underlying data for all analyses. All authors reviewed, revised, and approved the final manuscript.

IDO, LM, EAB, BOB, GL, GE, YJ, IM, and HATA, were supported by the Bill and Melinda Gates Foundation (Investment ID: INV-036449) and CHOIR.

## Conflict of interests

All authors declare no conflicts of interest.

## Artificial Intelligence Declaration

ChatGPT and Claude were used to assist with language editing and manuscript preparation. The authors verified all outputs, performed all scientific analyses and interpretations independently, and accept full responsibility for the manuscript’s content.

## Supplementary Information

Additional file: Supplementary material. ChatMRPT development, study procedures, data collection instruments, and supplementary analyses.

## Notes

### Competing Interest Statement

The authors have declared no competing interest.

### Author Declarations

Ethical approval for the survey was granted by the Institutional Review Board at Loyola University Chicago Health Sciences Division (Approval Number: 291901 and 218561) and authorization on transcription usage was obtained retrospectively from the Institutional Review Board at Loyola University Chicago Health Sciences Division (Approval Number: 219385) to cover transcribed audio recordings for phase one.

