## Supplementary material for "Institutionalizing LLM-assisted decision support for malaria risk-focused ITN reprioritization in Nigeria: Digital competency, workplace resource profiles, experiences, and pathways to routine integration"

### Introduction

This appendix provides supplementary information supporting the development, evaluation, and institutionalization assessment of ChatMRPT. Materials include details on MRPT development, participant characteristics, workplace-resource assessments, representative outputs from MRPT and ChatMRPT, and data-collection instruments used during stakeholder engagement and evaluation.

### Evolution timeline of MRPT to ChatMRPT

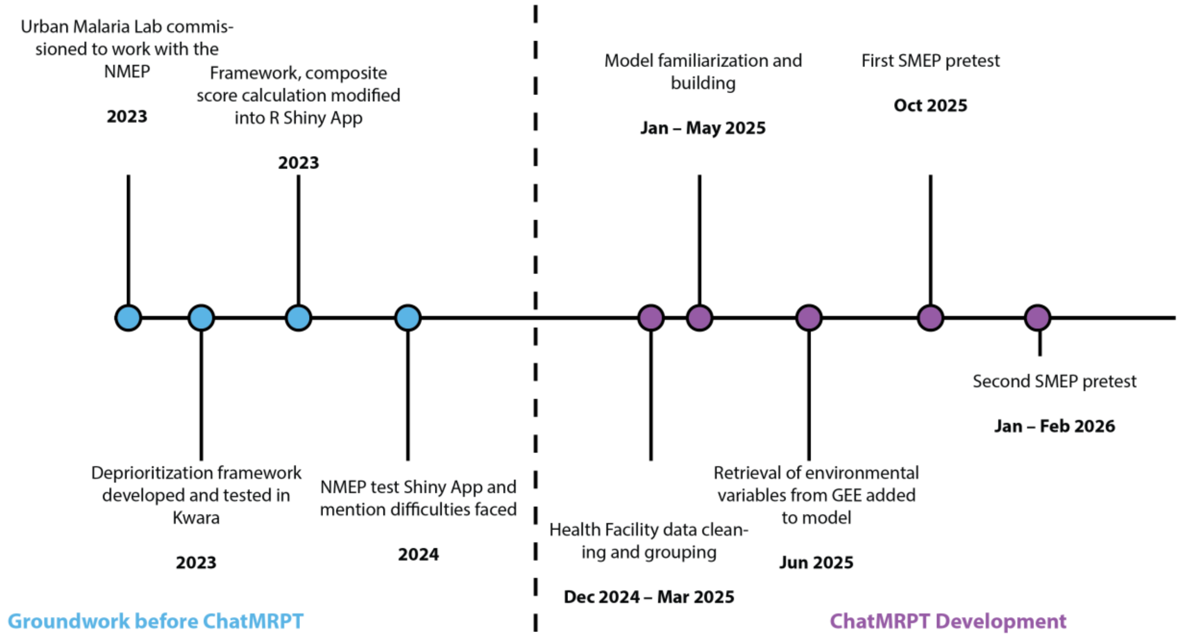

**Figure A1. Timeline of MRPT groundwork and ChatMRPT development.** The timeline summarizes key activities preceding and informing the development of ChatMRPT, from initial development and testing of the MRPT framework through model development, data integration, and testing with State Malaria Elimination Programme (SMEP) officers.

**Technical framework of developing MRPT.**

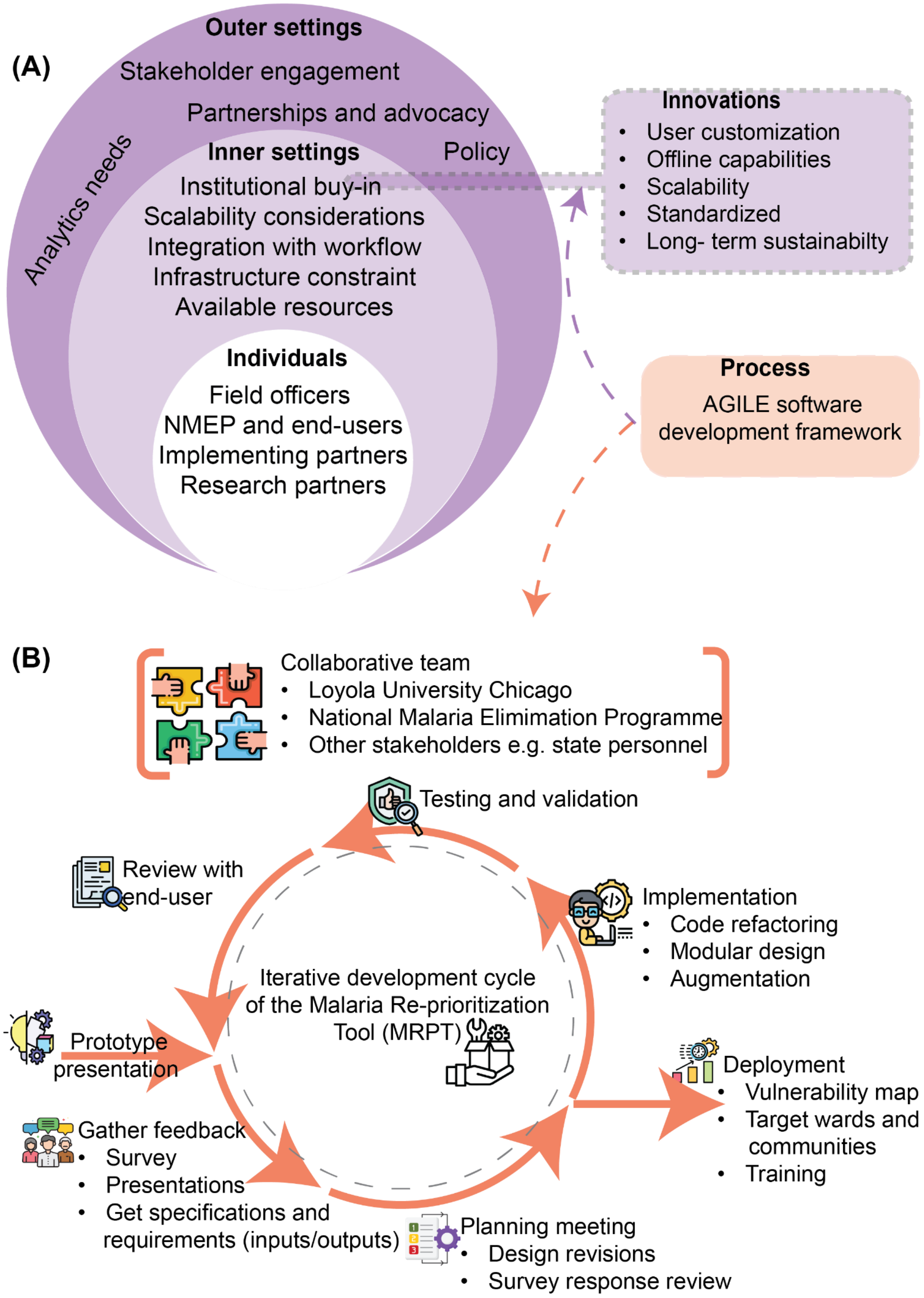

**Figure A2: Implementation and software-development frameworks.**

Figure A2 illustrates the implementation and software-development frameworks that informed the co-development of MRPT. The development process was guided by the Consolidated Framework for Implementation Research (CFIR) (1,2) and an Agile software development approach. CFIR informed consideration of contextual, organizational, and user-level factors, while Agile development (3–6) facilitated iterative refinement through stakeholder feedback, prototype testing, validation, and deployment activities. Together, these frameworks supported the development of a flexible, user-centered malaria reprioritization platform aligned with operational planning needs.

The CFIR framework informed consideration of outer-setting factors (policy, partnerships, stakeholder engagement), inner-setting factors (institutional buy-in, resources, workflow integration), individual stakeholders (NMEP, implementing partners, field officers, and researchers), innovation characteristics (customization, scalability, sustainability), and implementation processes. These considerations were operationalized through an Agile development cycle involving stakeholder engagement, prototype evaluation, iterative refinement, testing, and deployment.

**Underpinnings of ChatMRPT**

ChatMRPT is a conversational LLM-assisted decision support platform that helps malaria control teams analyze surveillance data and plan resource distribution at the ward level in Nigeria. Users type requests in plain language into a chat interface; the system, powered by OpenAI's GPT-4o model, interprets those requests and runs the appropriate analysis automatically (*See Figure A3*).

The workflow begins with the upload of ward-level surveillance data in a standardized spreadsheet format. Using built-in population estimates and administrative boundary layers, ChatMRPT calculates malaria burden metrics for each ward. The platform then integrates satellite-derived environmental datasets stored on the server, including rainfall, vegetation indices, nighttime light intensity, flood risk, and housing characteristics. Environmental variables were extracted at the ward level and aligned with Nigeria's geopolitical zones, reflecting regional differences in malaria risk drivers.

Malaria risk is assessed using two complementary approaches. The first generates a composite risk score by averaging normalized malaria burden and environmental indicators across selected variable combinations. The second applies Principal Component Analysis (PCA) to identify dominant patterns of variation across all variables without predefined weighting. Both approaches classify wards into High-, Medium-, and Low-risk categories.

The platform also incorporates a settlement classification module to support targeted intervention planning. Users review satellite imagery overlaid with a spatial grid and manually classify grid cells as Formal, Informal, Slum, or Rural settlements.

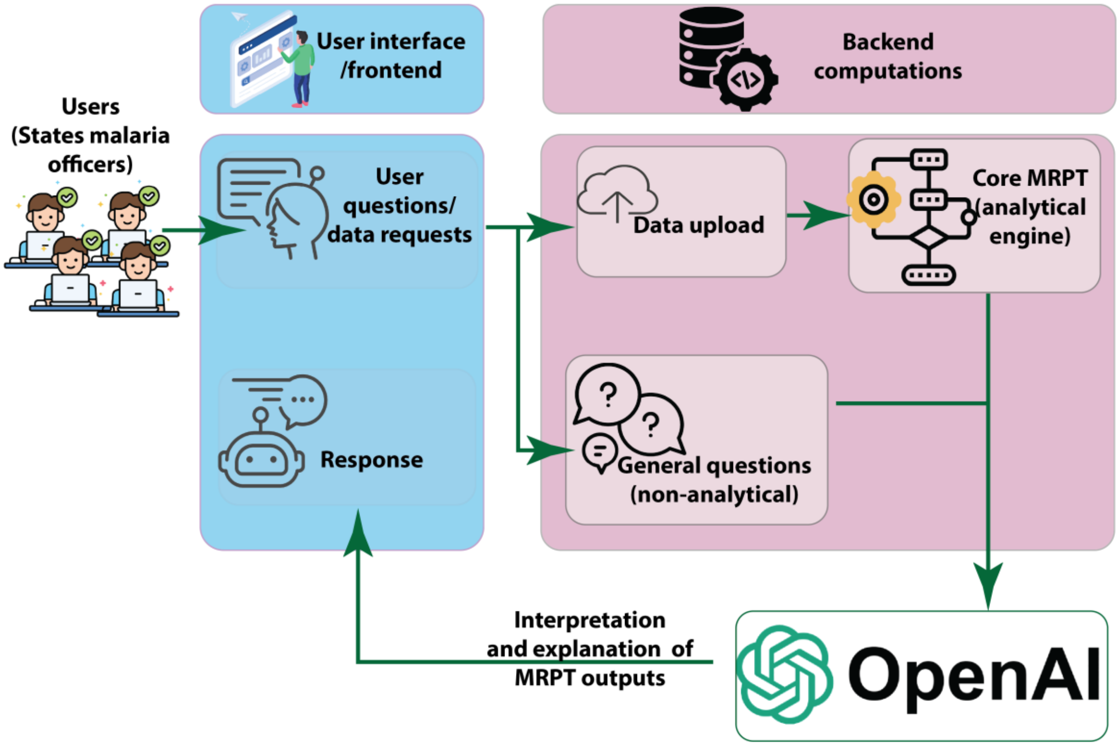

**Figure A3: A schematic diagram of ChatMRPT.**

Results are presented through interactive maps, charts, and summary tables. For any visualization, users can select the **Explain** function to obtain an AI-generated plain-language interpretation of the results. Final outputs include ward- and LGA-level datasets, settlement classification maps, malaria risk maps, and ITN allocation plans based on ward-level risk and population estimates. All outputs can be exported and downloaded as a consolidated analysis package.

**<u>Phase Two</u>**

The study included participants from 28 of Nigeria's 37 states (including FCT). Figure A4 shows their geographic distribution and the number of participants from each state.

**Study area**

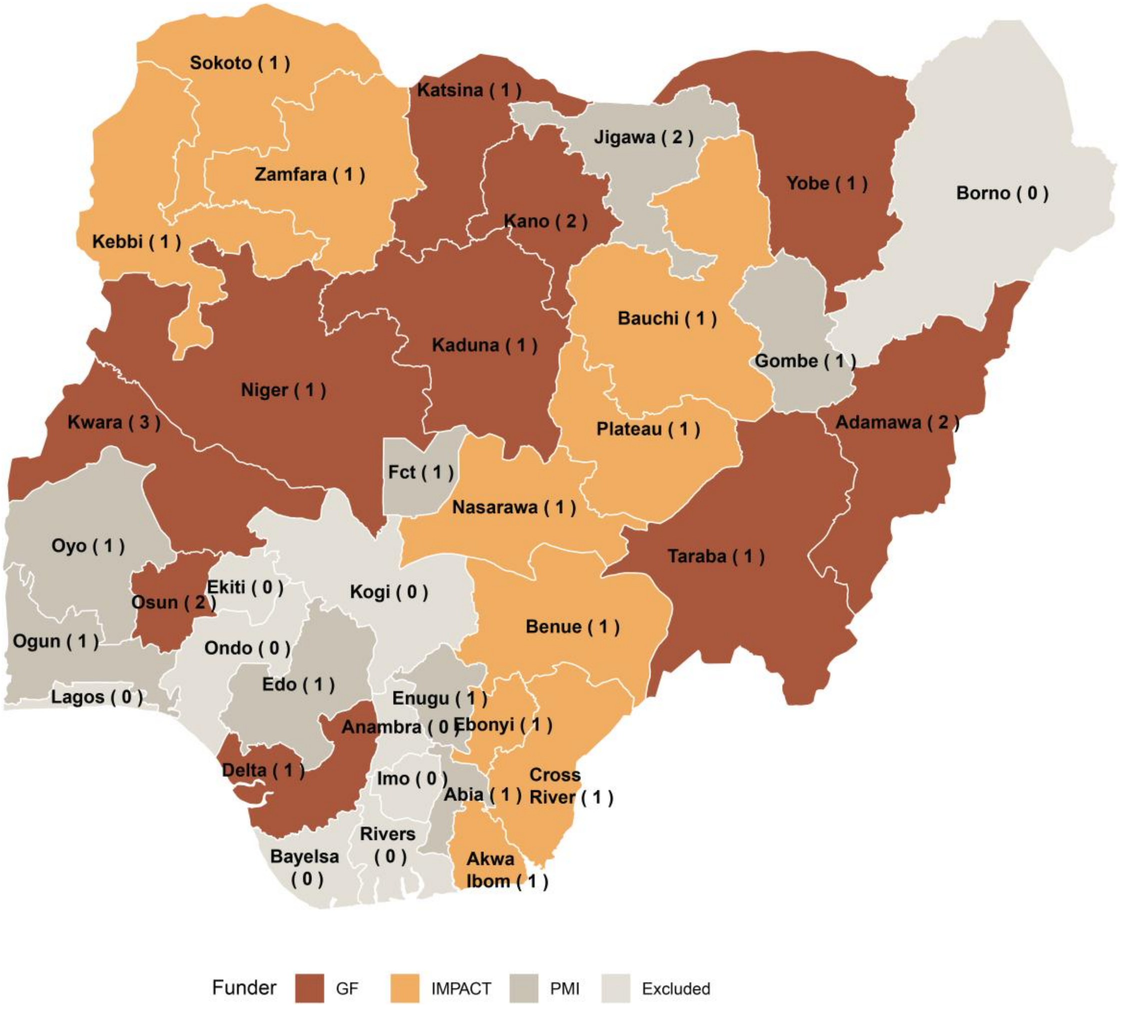

**Figure A4: Study area map**

**Highlights the study area showing the total number of participants**

*Functional categories of digital tools*

Participants were asked to report digital tools they routinely used in their work. Reported tools were grouped into seven functional categories: GIS and mapping tools (e.g., QGIS, ArcGIS, Google Earth), data collection platforms (e.g., KoboToolbox, ODK, SurveyCTO), dashboards and visualization tools (e.g., Power BI, Tableau), health information systems (e.g., DHIS2, NMDR, DiGiT), analytical software (e.g., Excel, R), AI-assisted tools (e.g., ChatGPT, DeepSeek), and malaria program-specific tools (e.g., Red Rose). Responses that did not fit these categories were classified as "Other." The responses were presented on a bar chart.

**Educational, Professional Experience and Job Rsponsibilities (Phase Two)**

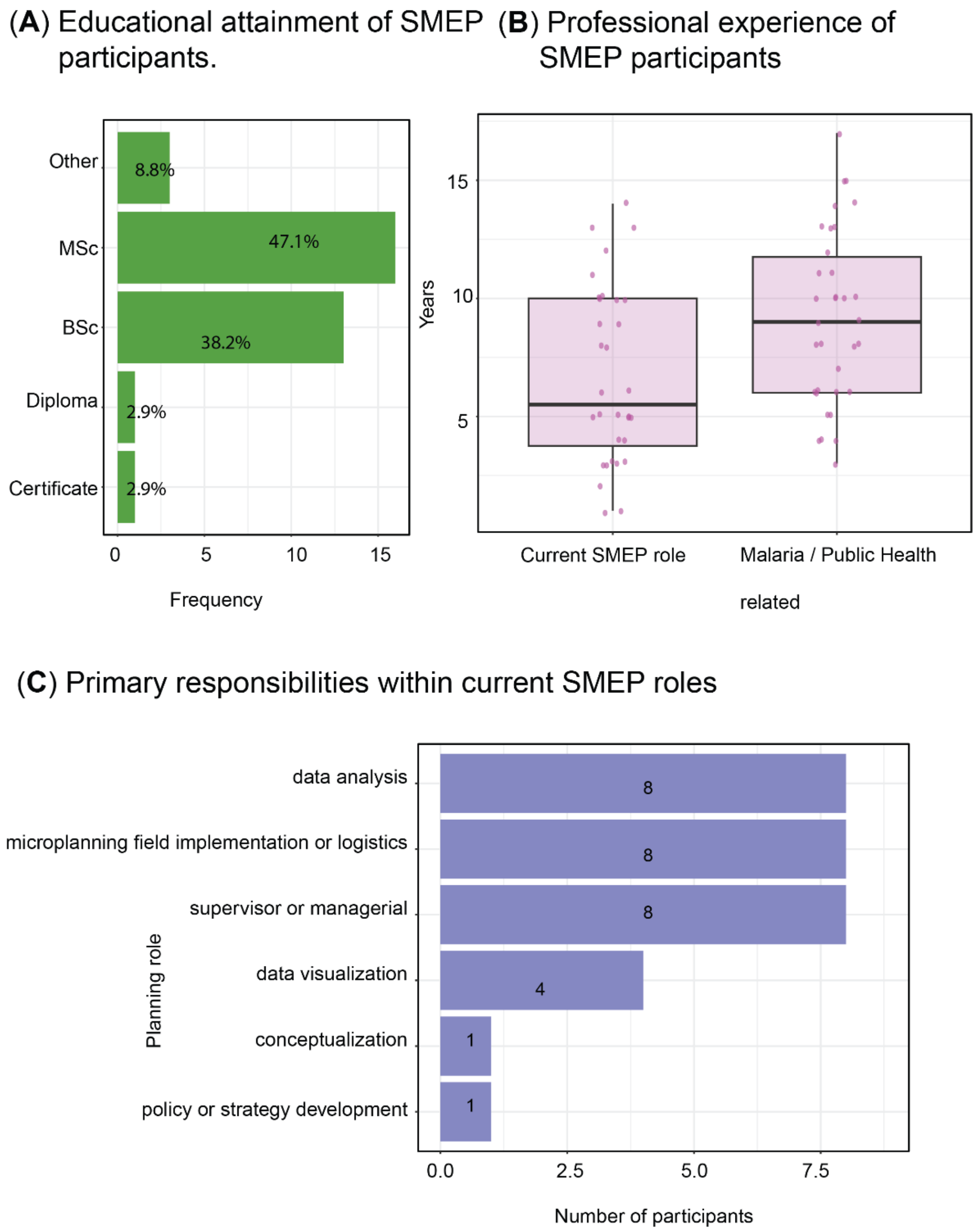

**Figure A5. Educational background, professional experience, and planning responsibilities of SMEP participants involved in the ChatMRPT evaluation.** *(A) Educational attainment of participating SMEP personnel. (B) Years of experience in current SMEP roles and in malaria/public health-related activities. (C) Self-reported responsibilities within current roles.*

*Participants were predominantly Masters- and bachelor's-level professionals, reported substantial experience in malaria and public health activities, and occupied roles spanning program management, microplanning, logistics, data analysis, and data visualization.*

The SMEP participants represented experienced malaria-program personnel with responsibilities directly related to malaria planning, implementation, monitoring, and decision-making (Appendix Figure A5). Most participants held Masters- or bachelor's-level qualifications (85.3%), reflecting a workforce with formal training relevant to public health and program implementation. Participants also reported substantial experience in both their current SMEP positions (*median of 6 years*) and broader malaria/public health activities (*median of 8 years*), indicating familiarity with routine malaria control operations and planning processes.

Primary responsibilities included program supervision and management, microplanning and logistics, data analysis, and data visualization, and each participants reported their major responsibility. These roles are directly involved in the interpretation of epidemiological data, intervention planning, resource allocation, and implementation oversight. Together with the digital readiness and workplace-resource assessments presented in the main manuscript, these findings suggest that participants possessed both the domain expertise and operational experience necessary to provide informed feedback on the usability, interpretability, and potential integration of ChatMRPT within routine malaria planning workflows.

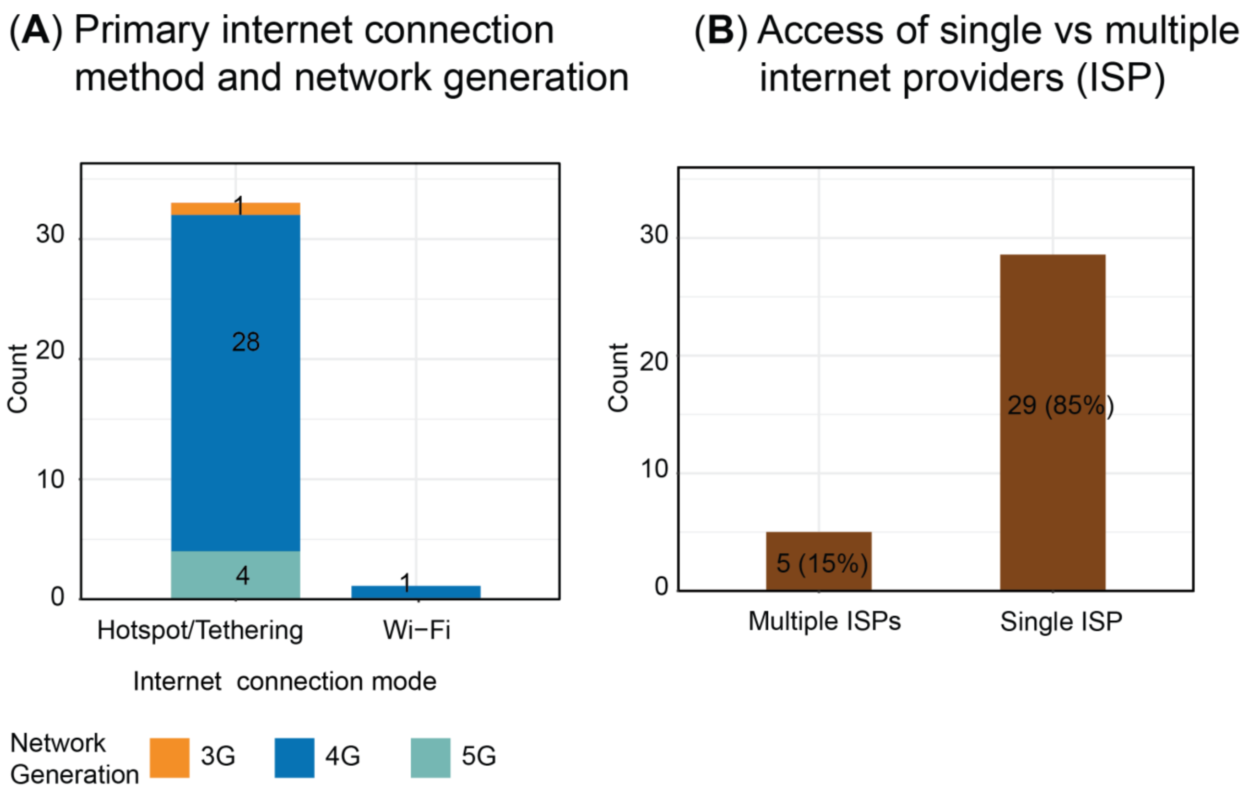

**Figure A6. Technology and connectivity resources used by SMEP participants during the pretest.** *(A) Internet connection methods and network generations. (B) Internet service providers used during the pretest. (C) Computer brands used.*

During the pretest most participants relied on mobile hotspot/tethering connections for internet access (33/34, 97%), with only one participant reporting having been connected through Wi-Fi. Among those using mobile connectivity, fourth-*generation* (4G) networks were predominant, while only a small proportion reported access to fifth-generation (5G) services. Participants also reported limited connectivity redundancy, with 86% (29/34) relying on a single internet provider and only 14% (5/34) having access to multiple providers. These findings provide important context for interpreting workplace-resource readiness. Although participants generally possessed the skills, experience, and digital competencies required to engage with ChatMRPT, access to reliable connectivity appeared more constrained. During the pretest, several participants experienced internet-related disruptions, including temporary disconnections and the need to rejoin sessions, highlighting the practical challenges of using web-based decision-support systems in routine work environments. Given that many malaria-program activities are sometimes conducted remotely or in field settings where connectivity is less stable, these findings suggest that infrastructural limitations, rather than individual capability, may represent a more immediate barrier to routine adoption. This observation reinforces the broader findings of the study that successful implementation of AI-supported planning tools depends on user readiness and on the availability of reliable digital infrastructure to support their use.

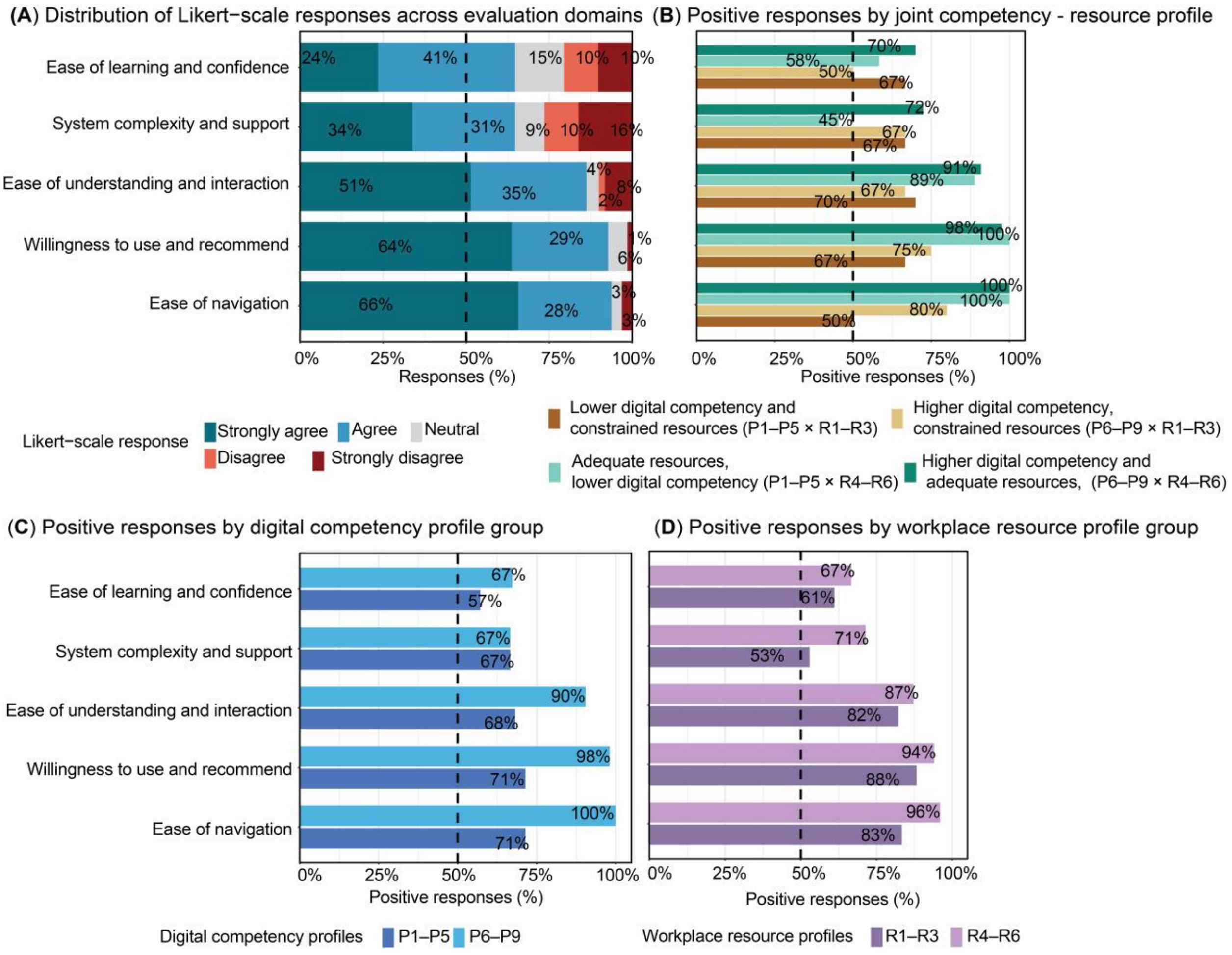

**Figure A7. Response-level perceptions of ChatMRPT across evaluation domains and participant profiles.** *(A) Distribution of Likert-scale responses across the five evaluation domains. (B) Positive responses (agree or strongly agree, after reverse-coding negatively worded items) by joint digital competency–workplace resource profile. (C) Positive responses by digital competency profile (P1–P5 vs P6–P9). (D) Positive responses by workplace resource profile (R1–R3 vs R4–R6). Percentages represent responses across questions grouped within each domain rather than participant-level domain averages. Dashed lines indicate 50%.*

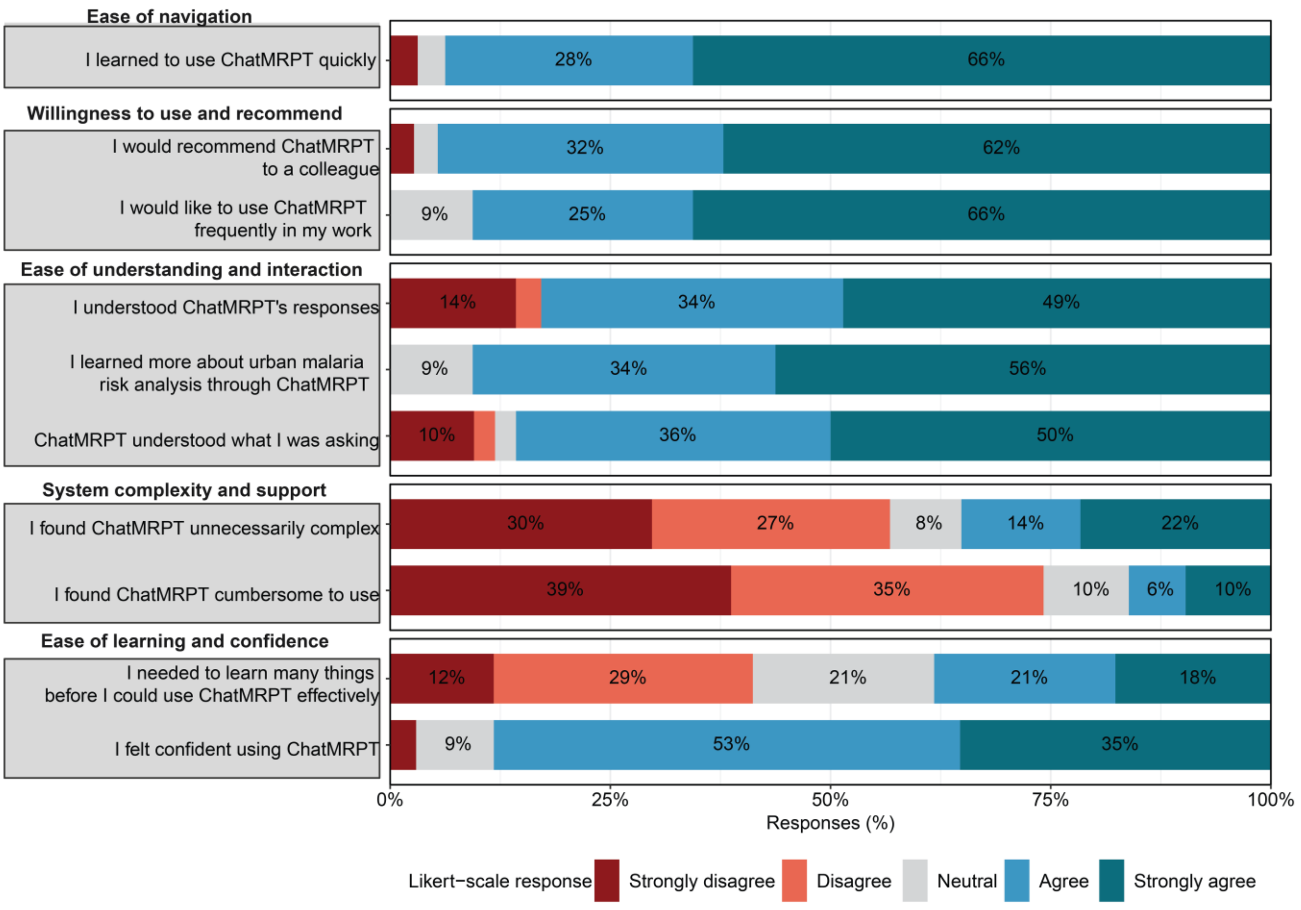

**Figure A8. Distribution of Likert-scale responses to individual ChatMRPT evaluation questions.** *Responses to individual survey questions are presented within the five conceptually related evaluation domains used in the participant-level analysis. Negatively worded questions are displayed as originally administered; responses were reverse-coded only for analyses in which higher scores represented more favourable perceptions.*

Supplementary response-level analyses were broadly consistent with the participant-level patterns while revealing variation across questions within domains (Figures A7–A8). In particular, the lower participant-level positivity for ease of learning and confidence reflected mixed responses regarding the amount participants needed to learn to use ChatMRPT effectively, despite most participants reporting confidence in using the platform. Item-level responses also showed that participants generally disagreed that ChatMRPT was cumbersome, while perceptions of unnecessary complexity were more varied.

**Appendix Two: ChatMRPT Standard Workflow — Prompts Start to Finish**

- Step 1: Start the workflow
  - *start the tpr workflow*
- Step 2: Select the state
  - *Type the state name, e.g. Kwara*
  - *or the number from the list shown*
- Step 3: Select facility level
  - *primary*
  - *secondary*
  - *tertiary*
  - *All*
- Step 4: Select age group
  - *under 5*
  - *pregnant women*
  - *above 15*
  - *all ages*
- Step 5: Map the variables
  - *plot the variable maps*
  - *or map [variable name] e.g. map rainfall*
- Step 6: Run the risk analysis
  - *run the malaria risk analysis*
- Step 7: View vulnerability maps
  - *show the vulnerability map*
  - *For PCA specifically: show the vulnerability map using PCA*
- Step 8: ITN distribution planning
  - *plan ITN distribution*
  - *(system will ask for total nets and average household size if not provided)*
- Step 9: Settlement classification
  - *create settlement classification*
  - *For top-risk wards: create settlement classification for the top 5 high risk wards*

- Example interrogation questions (interpretation, educational, user exploration, anything else)
    - *What is urban microstratification*
    - *What are the variables important to consider in investigating the risk of malaria in a state*
    - *What is NDMI and how does it influence the transmission of malaria*

**Appendix Three: Tools/Instruments used in the data collection for the MRPT ChatMRPT.**

**Instrument One:** Malaria Risk Mapping Tool (MRMT)Software Application - Feedback Survey

The MRMT (Malaria Risk Mapping Tool) app is designed to enhance malaria control efforts in urban areas by providing key features such as risk mapping, identification of low/high malaria risk areas at the ward level to ensure resource allocation to at-risk communities and de-prioritization of low-risk areas. We aim to make the app user-friendly and tailored to your needs.

We are conducting a survey to better understand and prioritize the features that matter most to you in the MRMT app. Your feedback will help us fine-tune the app to meet the needs of the National Malaria Elimination Program (NMEP) and other users.

Please take 35 minutes to evaluate the app's features, considering both usability and functionality. Your participation is voluntary, and we value your honest feedback.

**To demo the software application, click here to access the app. Click this link to access the demo data and shapefile.**

**Part 1: General Information**

1. Name of your organization? * (*Mark only one.*)
   - ❑ National Malaria Elimination Programme (NMEP)
   - ❑ State Ministry of Health (SMEP)
   - ❑ Federal Ministry of Health (FMOH)
   - ❑ State Malaria Elimination Program (SMOH)
   - ❑ Catholic Relief Services (CRS)
   - ❑ World Health Organization (WHO)
   - ❑ Other ______________________________
2. What is your role within your organization? *(*Mark only one.)*
   - ❑ Programme Officer
   - ❑ Programme Manager
   - ❑ Statistician
   - ❑ Data Analyst/Scientist
   - ❑ Project Officer
   - ❑ Field Health Worker

- ❑ Other ______________________________

3. How good are you at using, navigating, and handling web applications? **(Mark only one.)*
   - ❑ Beginner
   - ❑ Intermediate
   - ❑ Advanced
   - ❑ Expert
4. How frequently do you use data visualization tools or web applications to analyze data? * *(Mark only one.)*
   - ❑ Daily
   - ❑ Weekly
   - ❑ Monthly
   - ❑ Rarely
   - ❑ Never

**Part 2: Experience with urban microstratification**

5. What is your job description? *
6. What are the daily tasks associated with your job? *
7. Have you ever been involved in urban microstratification or de-prioritization? *
   - ❑ Yes
   - ❑ No
8. If yes, what methods were implemented? *(Select all that apply)*
   - ❑ GIS software
   - ❑ Statistical software
   - ❑ Household/ward-level assessment
   - ❑ Contracted experts
   - ❑ Mathematical models
   - ❑ Remote classification
   - ❑ Other ______________________________
9. What variables did you consider? *(Select all that apply)*
   - ❑ Rainfall

- ❑ Distance to water
- ❑ Temperature
- ❑ Vegetation index
- ❑ Floods
- ❑ Slums
- ❑ Precipitation
- ❑ Test positivity rate
- ❑ Humidity
- ❑ Housing quality
- ❑ Dumpsites
- ❑ DHS prevalence
- ❑ Other ______________________________

10. Where do you obtain data? (Select all that apply)
    - ❑ DHIS2
    - ❑ NMDR
    - ❑ Government databases
    - ❑ Academic institutions
    - ❑ Crowdsourced data
    - ❑ Private sector
    - ❑ International organizations
    - ❑ Other ______________________________
11. What challenges have you encountered?
12. What solutions have helped?

**Part 3: App features and usability**

13. Does MRMT address your challenges? *
    - ❑ Yes
    - ❑ No
    - ❑ Not sure
14. Please explain your answer.
15. Who is the end user, and would you use the app?

**Part 4: Decision-Making and Output Interpretation**

16. Rate the following:
    - ❑ Easy to navigate
    - ❑ Clear instructions
    - ❑ Easy data upload
17. Does the app help decision-making?
    - ❑ Yes
    - ❑ Neutral
    - ❑ No

**Part 5: Additional Feedback**

18. Would you prefer more guidance?
    - ❑ Yes
    - ❑ Neutral
    - ❑ No
19. Which outputs are helpful? (Select all that apply)
    - ❑ Distribution plots
    - ❑ Composite scores
    - ❑ Box plots
    - ❑ Normalization plots
    - ❑ Decision tree
20. Would you use the application in the future?
    - ❑ Yes
    - ❑ No
21. Explain your answer.
22. What alternatives do you use?
23. Share your experience.
24. Additional comments.

**Instrument Two:** ChatMalaria Risk Mapping Tool (MRPT) Software Application – pre and post pretest survey

**To build a clear profile of participants'capacities, contexts, and potential needs for adopting MRPT/chatMRPT) (*To help us track responses anonymously, please enter the last four digits of your phone number. This will not be used to identify you personally.*)**

**Study Title:** A Mixed-Methods Evaluation of chatMRPT: an AI-Enhanced Copilot for Malaria Reprioritization in Urban Nigeria.

Background information

1. What is your age?

   (18 – 24) / (25 – 34) / (35 – 44) / (45 – 54) / 55+

2. What is the highest level of education you have completed?

   Primary education/Secondary education / Diploma / Certificate/ Bachelor's degree/ Master's degree/ Doctorate (PhD/DrPH/MD)/Others(specify)__________

3. How many total years of formal education have you completed? (___ years)
4. How many years have you worked in your current role? (___ years)
5. How many total years have you worked in malaria control or public health? (___ years)
6. What is your current job title/position? (___________________)
7. At which level do you primarily work?
8. Have you previously used digital decision-support tools (e.g., GIS platforms, dashboards, or AI tools)?

   Yes/No

   If yes, please specify which tools: (Open response)

9. What type of training/learning format do you learn best from (hands on workshop/ demonstration/self-paced manuals/videos/ peer to peer learning/ other (specify))

Technical and digital literacy

1. How would you rate your computer skills? (beginner/Intermediate/advanced)
2. How often do you work with data in your role (e.g., analysis, interpretation, reporting)? (rarely/sometimes/often/very often)
3. Do you have regular access to the following resources while at work? (laptop or desktop/smartphone or tablet/ stable internet/ stable electricity) *tick all that applies*

Decision making and planning

1. How frequently are you directly involved in ITN campaign planning or microstratification activities? (rarely/sometimes/often/very often)
2. What role do you usually play in planning processes (data analysis/conceptualization/data visualization/microplanning field implementation or logistics/ supervisor or managerial/ policy or strategy development/ other (*specify*))

**Instrument Three: Likert ChatMRPT Usability Questions**

1. Understanding and Interaction
    a. ChatMRPT understood what I was asking
        Type: Scale 1–5 (Strongly disagree → Strongly agree)
    b. I understood ChatMRPT's responses"
        Type: Scale 1–5 (Strongly disagree → Strongly agree)
    c. I learned more about urban malaria risk analysis through ChatMRPT
        Type: Scale 1–5 (Strongly disagree → Strongly agree)
2. Ease of Use
    a. I learned to use ChatMRPT quickly"
        Type: Scale 1–5
3. Recommendation and Engagement
    a. I would recommend ChatMRPT to a colleague
        Type: Scale 1–5
    b. I would like to use ChatMRPT frequently in my work
        Type: Scale 1–5
4. System Complexity and Support
    a. I found ChatMRPT unnecessarily complex
        Type: Scale 1–5

b. I found ChatMRPT cumbersome to use

   Type: Scale 1–5

5. Confidence and Learning Curve
   a. I felt confident using ChatMRPT

      Type: Scale 1–5

   b. I needed to learn many things before I could use ChatMRPT effectively

      Type: Scale 1

6. Open Feedback
   a. Please share your thoughts about ChatMRPT. What did you like? What could be improved?
   b. What did you think of ChatMRPT's responses? Were they clear and helpful, or confusing?

**<u>Instrument Four:  Focus Group Discussions two (Perspectives on chatMRPT)</u>**

A Mixed-Methods Evaluation of chatMRPT: an AI-Enhanced Copilot for Malaria Reprioritization in Urban Nigeria.

**Purpose**

To evaluate users' final perceptions and experiences with chatMRPT before/after completing the training, (see Appendix 4 for learning objectives), and post-training use phase. The discussion will assess acceptance and reveal opportunities for improvement based on predefined domains and five key dimensions: usability, interpretability, human–computer interaction, functionality, and complexity.

Participants will share qualitative insights on these five dimensions after independently exploring chatMRPT. A shared record (e.g., Google Docs or a structured live-notes template) will be maintained for all group sessions. Comments should be tagged by theme: Usability (U), Interpretability (I), Functionality (F), Complexity (C), and Human–Computer Interaction (HCI), and Value (V), to enable rapid thematic analysis.

**Preamble**

***Good afternoon, everyone.***

*Thank you for joining this post-training discussion. My name is [Facilitator's Name], and I'm part of the Urban Malaria Lab team. Over the past 4 days you've been introduced to the chatMRPT tool, explored its features, and applied it to real-world malaria microplanning scenarios using your own state data.*

*This session is an opportunity to reflect on your experience now that you've had hands-on exposure. We want to hear about what worked well, where you encountered challenges, and how the tool fit or didn't fit into your planning processes. We're also interested in your impressions of its usability, interpretability, complexity, functionality, the quality of human–computer interaction, and its value add.*

*Your feedback will help us understand how chatMRPT performs in practice, what changes or enhancements are needed, and how it might be best integrated into ongoing decision-making for urban malaria intervention planning and reprioritization.*

*There are no right or wrong answers. Please be candid—even if something was frustrating or confusing. We're here to learn from your experience, and everything you share will remain confidential, used only to improve the tool and for analysis in related research outputs.*

*We'll guide the discussion with structured questions, but you're encouraged to share examples, compare experiences, and build on one another's points. Everyone's voice matters, and we'll make sure all perspectives are heard.*

*With that, let's begin…*

***Discussion Questions by Theme***

1. **Describe your overall experience with regards to using ChatMRPT?**
    - *What stood out most during your interaction with ChatMRPT?*
        - Was what stood out mostly positive, negative, or unexpected?
        - Were there particular features, responses, or moments that stayed with you?
        - Did anything differ from what you expected before using the tool?
    - *What felt intuitive or easy to pick up?*
        - Was navigation through the interface straightforward?

        - Were there parts where you knew what to do next without guidance?
        - Did prior experience with similar tools help or not?
    - *What felt confusing or slowed you down?*
        - Was the difficulty related to wording, explanations, or navigation?
        - Were there moments where you had to stop and think about what to do next?
        - Did any uncertainty persist even after continued use?
    - **If you had to describe your overall experience using ChatMRPT in one or two words, what would they be?**

2. **How did interacting with ChatMRPT shape your understanding of how risk-based reprioritization is done?**
    - *How did this compare to how you usually understand or work with prioritization outputs?*
        - Did ChatMRPT explain things differently from the tools or reports you currently use?
        - Were there aspects that felt more familiar or unfamiliar?
        - Did the explanations help you understand *why* certain areas were prioritized?
        - Were there parts of the process that became clearer - or still unclear?
        - Did the way results were presented match how you think about malaria risk?
    - How relevant were the explanations and maps for the decisions you are typically involved in?
        - Did they help you move from seeing results to thinking about action?
        - Were there decisions where the outputs felt less applicable?
    - What aspects of ChatMRPT were still difficult to interpret or use?
        - Were there explanations or outputs that raised more questions than they answered?
        - Where did you still feel unsure or hesitant?
    - Did ChatMRPT address any challenges you normally face when interpreting prioritization results?
    - Were there moments where it made your task easier or clearer than usual?
        - Which parts of your usual workflow did it help with most?

    - Did this affect how quickly you could move from data to decisions?
- Is the explanation on how the results are generated understandable and sufficient?

3. **How does ChatMRPT fit into how routine planning decisions (ITNs campaigns) are normally made in your state?**
    - At what stage of planning would ChatMRPT realistically be used?
        - How would it typically be used at that stage?
        - Did it feel like a natural part of the process or an additional step?
    - How did ChatMRPT relate to the tools and data systems you already use (e.g., DHIS2, Ipolongo, Red Rose)?
        - Did it feel complementary, duplicative, or conflicting?
        - Were there moments where the logic or outputs did not align with your usual processes?
        - How did you respond to those moments did you correct or nudge it
    - What integrations or changes would increase trust or efficiency in routine use?
4. **After using ChatMRPT, how confident do you feel explaining the prioritization results to others?**
    - Who did you feel confident explaining the results to (e.g., peers, supervisors, partners)?
        - Were there audiences you felt less comfortable explaining them to? Why?
    - What explanations, narratives, or visual elements helped most when explaining the results
        - Were there parts of ChatMRPT that helped you explain why areas were prioritized?
    - What aspects remained difficult to explain or defend?
        - What questions on reprioritization do you anticipate will be difficult to answer using chatMRPT?
5. **What would need to happen for ChatMRPT to move from a helpful tool to something your institution expects or encourages staff to use?**
    - What capacities are most important?
    - Data availability and quality?
    - Staff skills?

    - Leadership or political backing? Who would need to endorse or approve its use?
- What evidence or assurances would be required?
- What barriers would still need to be addressed?

6. **Did using ChatMRPT feel like a safe way to support decision-making in your institutional context**
    - In what situations did it feel safe or unsafe to rely on ChatMRPT?
    - What risks; technical, professional, or political; do you still perceive?
    - What would make it feel more legitimate or official?
7. **How could ChatMRPT be better embedded into routine planning and integrated with other tools?**
    - How could chatMRPT strengthen decision-making at higher levels (state or national), not just in your daily work?
    - While using the tool, were new concepts explained in a way that you could comprehend? Can you give an example which aided your comprehension?
    - What aspects or concepts do you think will feel complex or inaccessible to some users?
8. **Please share up to three suggestions that would make ChatMRPT more useful or easier to institutionalize.**
    - What organizational, technical, or policy changes would be required?
    - What training or capacity-building would staff need?
    - How should ChatMRPT fit within existing malaria planning and decision-making workflows?
    - Who would need to support or approve its routine use?
    - What challenges/barriers might prevent long-term adoption?
